# Towards patient-relevant, trial-ready digital motor outcomes for SPG7: a cross-sectional prospective multi-center study (PROSPAX)

**DOI:** 10.1101/2024.01.09.24301064

**Authors:** Lukas Beichert, Jens Seemann, Christoph Kessler, Andreas Traschütz, Doreen Müller, Katrin Dillmann-Jehn, Ivana Ricca, Sara Satolli, Ayşe Nazli Başak, Giulia Coarelli, Dagmar Timmann, Cynthia Gagnon, Bart P. van de Warrenburg, PROSPAX consortium, Winfried Ilg, Matthis Synofzik, Rebecca Schüle

## Abstract

**Background and Objectives:** With targeted treatment trials on the horizon, identification of sensitive and valid outcome measures becomes a priority for the >100 spastic ataxias. Digital-motor measures, assessed by wearable sensors, are prime outcome candidates for SPG7 and other spastic ataxias. We here aimed to identify candidate digital-motor outcomes for SPG7 – as one of the most common spastic ataxias – that: (i) reflect patient-relevant health aspects, even in mild, trial-relevant disease stages; (ii) are suitable for a multi-center setting; and (iii) assess mobility also during uninstructed walking simulating real-life.

**Methods:** Cross-sectional multi-center study (7 centers, 6 countries). Unaided walking was assessed in 65 patients with SPG7 and 50 unrelated healthy controls using 3 wearable sensors (Opal APDM). Digital gait measures were correlated to measures of disease severity (SARA, SPRS; including mobility-relevant subscores SPRS^1315291292025^, SARA^PG^) and activities of daily living (FARS-ADL). The task set included lab-based defined gait tasks, complemented by uninstructed ‘supervised free walking’.

**Results:** Among 30 hypothesis-based gait measures, 18 demonstrated at least moderate effect size (Cliff’s δ>0.5) in discriminating SPG7 patients from controls, and 17 even in mild disease stages (SPRS^mobility^≤9). Spatiotemporal variability measures such as the spatial variability composite measure SPcmp (ρ=0.67, p=<0.0001), Stride Time CV (ρ=0.67, p=<0.0001) and Swing CV (ρ=0.64, p=<0.0001) showed the highest correlations with clinician-reported mobility scores (SPRS^mobility^), and overall disease severity (SPRS, SARA). Overall, top-ranked measures also correlated with patient-relevant functional deficits in everyday life activities (FARS-ADL). In mild disease stages (SPRS^mobility^≤9, n=41), Swing CV (ρ=0.53, p=<0.0001) and SPcmp (ρ=0.50, p=<0.0001) correlated with SPRS^mobility^. In the uninstructed ‘supervised free walking’ task, the correlations between spatiotemporal variability measures (Stride Time CV, Stride Length CV, Swing CV) and SPRS^mobility^ could be confirmed; additionally, Gait Speed (ρ=-0.59, p=<0.0001) was highly correlated with SPRS^mobility^.

**Discussion:** We here identified trial-ready digital-motor candidate outcomes for the spastic ataxia SPG7, all characterized by proven multi-center applicability, ability to discriminate patients from controls, and correlation with measures of disease severity – even in mild disease stages –, and patient-relevant everyday function. If validated longitudinally, these sensor outcomes might inform future natural history and treatment trials in SPG7 and other spastic ataxias.

## Introduction

As we are entering the era of genomic therapies for rare diseases, the identification of reliable and valid outcome parameters with sensitivity to longitudinal change and treatment effects is becoming a priority task. Quantitative digital motor outcomes, assessed through body-worn sensors, could potentially meet this need by capturing changes in patient-relevant health aspects within trial-like time frames, even in moderately progressive diseases like Hereditary Spastic Paraplegia Type 7 (SPG7).

SPG7 is an autosomal recessive hereditary neurodegenerative disease, manifesting as spastic ataxia typically in adulthood (Casari & Marconi, 1993). Although currently no curative treatment exists, progress in the development of genomic therapies for genetic spastic ataxias, including anti-sense oligonucleotides and gene replacement strategies, and first treatment trials conducted in diseases like Spastic Paraplegia type 50 (SPG50, NCT05518188) or Spinocerebellar Ataxia Type 1 and 3 (SCA1 / SCA3, NCT05822908), raises hopes that potential therapeutic agents for SPG7 might become available in the near future.

Digital gait measures have demonstrated promising properties in hereditary ataxias, reliably discriminating between patients and controls, correlating cross-sectionally with clinical measures of disease severity and – in the few longitudinal studies conducted so far – also exhibiting sensitivity to capturing change (Ilg et al., 2020; Seemann et al., 2023; Shah et al., 2021; Thierfelder et al., 2022; Velazquez-Perez et al., 2021). In contrast, only few studies on sensor-based gait analysis have been conducted in hereditary spastic paraplegias (Loris et al., 2023; Regensburger et al., 2022), and no study has yet systematically employed body-worn sensors to examine gait in diseases like SPG7 or other spastic ataxias. Anticipated treatment trials in diseases like SPG7 will likely (i) focus on patients in early disease stages and (ii) involve multiple centers to reach sufficient power. Validation of sensor gait measures as potential treatment outcomes should therefore specifically assess performance in those early disease severity strata and demonstrate multi-center applicability. Moreover, for sensor gait measures to gain regulatory and patient acceptance as outcome parameters, they need to reflect health aspects that are relevant to patients ((FDA), 2023). Therefore, performance of sensor outcomes needs to be assessed in patient-relevant settings, i.e. conditions resembling patients’ everyday lives, and gait measures should demonstrate correlation with clinical measures capturing disease-related impairment on a functional, patient-relevant level.

This cross-sectional study presents candidate digital gait outcomes for spastic ataxias like SPG7, demonstrating discriminative power and correlation with clinical outcome assessments of patient-relevant health aspects. Evaluated across multiple centers, in gait assessments in settings simulating real-life, and with a special focus on patients in mild disease stages, the presented gait measures are prime candidate outcomes to be validated longitudinally and potentially be applied in future treatment trials.

## Methods

### Participants

The study cohort was part of the study “An integrated multimodal progression chart in spastic ataxias” (PROSPAX) funded by the European Union via the EJP-RD programme (ClinicalTrials.gov, No: NCT04297891). Seventy patients with genetically confirmed and clinically manifest SPG7 and 50 healthy controls (HC) with available gait recordings were recruited by seven centers in six countries based on the following inclusion criteria: (1) ability to walk at least 10m without walking aid; (2) absence of severe comorbidities (due to SPG7 or unrelated) which present a major confounder for evaluation of gait and stance such as: amputation, blindness, severe dementia, severe joint deformities or contractures, or fixed orthoses. HC had no history of any neurologic or psychiatric disease, no family history of neurodegenerative disease, and did not show any neurological signs upon clinical examination. After exclusion of invalid gait recordings (damaged data files, unreliable step detection) recordings of 65 SPG7 patients and 50 HC from the lab-based walking condition (LBW), and of 57 patients and 37 HC from the supervised free walking condition (SFW) remained suitable for analysis (supplementary figure 1). The Institutional Review Boards of all recruiting centers approved the study. All participants provided written informed consent before participation according to the Declaration of Helsinki.

### Clinical assessments

All participants underwent a detailed neurological examination. Disease severity was rated using the Scale for the Assessment and Rating of Ataxia (SARA) (Schmitz-Hubsch et al., 2006) and the Spastic Paraplegia Rating Scale (SPRS) (Schüle et al., 2006). Mobility-relevant SPRS items 1-6 were combined into a subscore termed SPRS^mobility^ (Gassner et al., 2021). SARA items 1-3 rating gait and posture were combined into the SARA posture & gait subscore (SARA^PG^) (Ilg et al., 2020; Lawerman et al., 2017). The Friedreich Ataxia Rating Scale/activities of daily living (FARS-ADL) was used to assess impact of the disease on patient-relevant health aspects and The Friedreich Ataxia Rating Scale Functional Staging (FARS Staging) was used to classify patients by function disease stages (Subramony et al., 2005).

### Gait conditions

Walking movements were recorded under two different conditions:

(1) Laboratory-based walking (LBW condition): Walking was constrained by a specified walking distance of 10 meters in a specific quiet non-public indoor floor and supervised by a study assessor watching the walking performance; participants were instructed to walk back and forth the walking distance at a self-selected speed; participants were asked to halt after 1 minute of walking and recordings were terminated.

(2) Supervised free walking (SFW condition): Largely unconstrained walking in public spaces in an institutional (hospital) compound (all indoor: 4 study sites; indoor and outdoor: 3 study sites), where participants were free to choose walking speed as they were lead along a predefined route lasting about 5-10 minutes; participants were accompanied by a study assessor watching the participant’s walking performance.

### Gait measures

Three inertial sensors (Opal, APDM Wearable Technologies Inc, Portland, WA) were attached on both feet and posterior trunk at the level of L5 with elastic Velcro® bands. Inertial sensor data were collected and wirelessly streamed to a laptop for automatic generation of gait and balance metrics by Mobility Lab software (APDM, Inc.). For the supervised free walking condition (SFW), data were logged on board of each Opal sensor and downloaded after the session. Step events and spatiotemporal gait features for each stride were extracted from the inertial measurement unit sensors using APDM’s *Mobility Lab* software (Version 2) (Mancini et al., 2011), which has been shown to deliver good to excellent accuracy and repeatability (Morris et al., 2019; Washabaugh et al., 2017). For the LBW condition, only recordings with a minimum number of 20 detected strides were included in the analysis. For the SFW condition, only strides from walking bouts of at least 5 consecutive strides were analysed.

We considered a hypothesis-based selection of 30 candidate gait measures based on previous studies and literature (from both the ataxia- and HSP-fields) and clinical plausibility (Buckley et al., 2018; Ilg et al., 2020; Laßmann et al., 2022; Martino et al., 2018; Regensburger et al., 2022; Shah et al., 2021; Velazquez-Perez et al., 2021). Gait measures for each recording were obtained by (i) computation of one or more of the following non-parametric measures for each of 14 of the gait features extracted for each stride by *Mobility Lab*: Median, normalized median absolute deviation (MADN = MAD/0.6745) and coefficient of variation (CV = median/MADN); (ii) computation of one composite measure of spatial step variability (SPcmp); and (iii) computation of median Harmonic Ratio (HR) of raw accelerometer signals from the lumbar sensor in 3 directions. A list of all gait measures and their definitions is provided in supplementary table 1.

The composite measure SPcmp was formed from Stride Length CV and Lateral Step Deviation to capture spatial step variability in both anterior-posterior and medio-lateral directions in one measure (Ilg et al., 2020). Briefly, the composite measure was determined for each individual participant in 2 steps: In step 1, the relative value of the participant in comparison to the value range of all participants was calculated for each of the 2 constituent measures separately (resulting in values between 0 and 1). In step 2, the greater of these 2 relative values was selected.

HR of pelvis linear acceleration was determined to quantify the smoothness of motion, as described previously (Ilg et al., 2020). Briefly, this method quantifies harmonic content of the acceleration signals in each direction (harmonic ratio anterior-posterior [AP], medio-lateral [ML], vertical [V]) using stride frequency as the fundamental frequency component. Using a finite Fourier series, HR is calculated by dividing the sum of the amplitudes of the first 10 even harmonics by the sum of the amplitudes of the first 10 odd harmonics for each given stride (Bellanca et al., 2013; Menz et al., 2003). A greater HR was interpreted as greater walking smoothness. HR measures have been shown to distinguish between patients with cerebellar disease and HCs under laboratory-based and real-life walking conditions (Ilg et al., 2020; Serrao et al., 2018).

### Statistics

For the hypothesis-based selection of 30 gait measures, the ability to discriminate between patients and controls was assessed by calculation of effect size Cliff’s δ (Cliff, 1996). Discriminative effect sizes were classified as *small* (δ ≥ 0.3), *moderate* (δ ≥ 0.5) or *large* (δ ≥ 0.8). Additionally, significance of group differences was determined by the non-parametric Wilcoxon signed-rank test. Group differences were considered significant when p < 0.05/n (n = 30: number gait measures), accounting for multiple comparisons.

For gait measures that discriminated patients from controls with at least moderate effect sizes, we assessed convergent validity by examining Spearman correlation between gait measures and 5 clinical outcome measures. The scales SPRS^mobility^ as a measure of mobility and SARA^PG^ as a measure of posture and gait were considered the most direct clinical equivalents to the sensor-based gait measures while at the same time reflecting health aspects of high relevance to patients and were thus treated as primary outcomes. As additional, exploratory outcomes, SPRS and SARA as standard measures of general disease severity in HSP and ataxia, respectively, and FARS-ADL as a measure of activities of daily living were included in the analysis. Effect sizes ρ are displayed with 95% confidence intervals (determined by boot strapping using MATLAB’s *bootci* function with 2000 samples) and p values. Effect sizes ρ were classified as *small* (ρ ≥ 0.1), *medium* (ρ ≥ 0.3), *large* (ρ ≥ 0.5), or very large (ρ ≥ 0.7)(Maher et al., 2013). Correlations between gait measures and primary clinical outcomes were reported as significant when p < 0.05/(n × m) (n: number of discriminative gait measures, m = 2: number of primary clinical outcome measures), hereby correcting for the number of comparisons. Correlations with exploratory outcomes SPRS, SARA and FARS-ADL were deemed as significant when p < 0.05. Further, spearman correlations between the two walking conditions (LBW, SFW) were calculated for each gait measure.

To evaluate the ability of the gait measures to discriminate mildly affected patients from controls, we performed a median split of the patient cohort with respect to SPRS^mobility^, thus defining a subgroup with mild (SPRS^mobility^ ≤ median_LBW_; termed ‘mild patient cohort’) and intermediate disease severity (SPRS^mobility^ > median_LBW_), where median_LBW_ denotes the median SPRS^mobility^ of all patients with valid recordings of LBW.

To assess whether gait measures differentiated between diseases stages defined by the FARS Staging, thus determining approximate meaningful score regions, patients were grouped into three stages (mild: ≤ 2.0, intermediate: 2.5-3.5, advanced: ≥ 4.0). Between-group/stage differences were analysed using the Kruskal-Wallis test, and – when the Kruskal-Wallis test yielded a significant effect – Wilcoxon ranksum test post hoc (considered significant when p<0.05). Statistical analysis was performed using MATLAB (version R2022a).

## Results

Gait recordings of 65 SPG7 patients and 50 HC from the LBW condition and of 57 patients and 37 HC from the SFW condition were analyzed. Individual participant characteristics are displayed in supplementary table 2. The majority of patients (61 of 65) exhibited clinical signs of both cerebellar and pyramidal systems involvement (supplementary figure 2).

### Measures of spatiotemporal gait variability discriminate between patients and controls with large effect sizes even in patients with mild disease severity

In LBW, the comparison between SPG7 patients and HC yielded large discriminative effect sizes of |Cliff’s δ| ≥ 0.8 for 5/30 gait measures, and at least moderate effect sizes of |δ| ≥ 0.5 for 18/30 gait measures. The strongest discrimination was observed for measures of spatial and temporal gait variability: SPcmp (δ = 0.90), Swing CV (δ = 0.86) and Lateral Step Deviation (δ = 0.84). (figure 1, table 1). Other measures displaying high discriminatory power included foot angle measures (Pitch at Initial Contact: δ = 0.81; Pitch at Toe Off: δ = 0.78) and measures of gait smoothness (Harmonic Ratio V: δ = -0.80; Harmonic Ratio AP: δ = -0.79).

**Figure 1:**
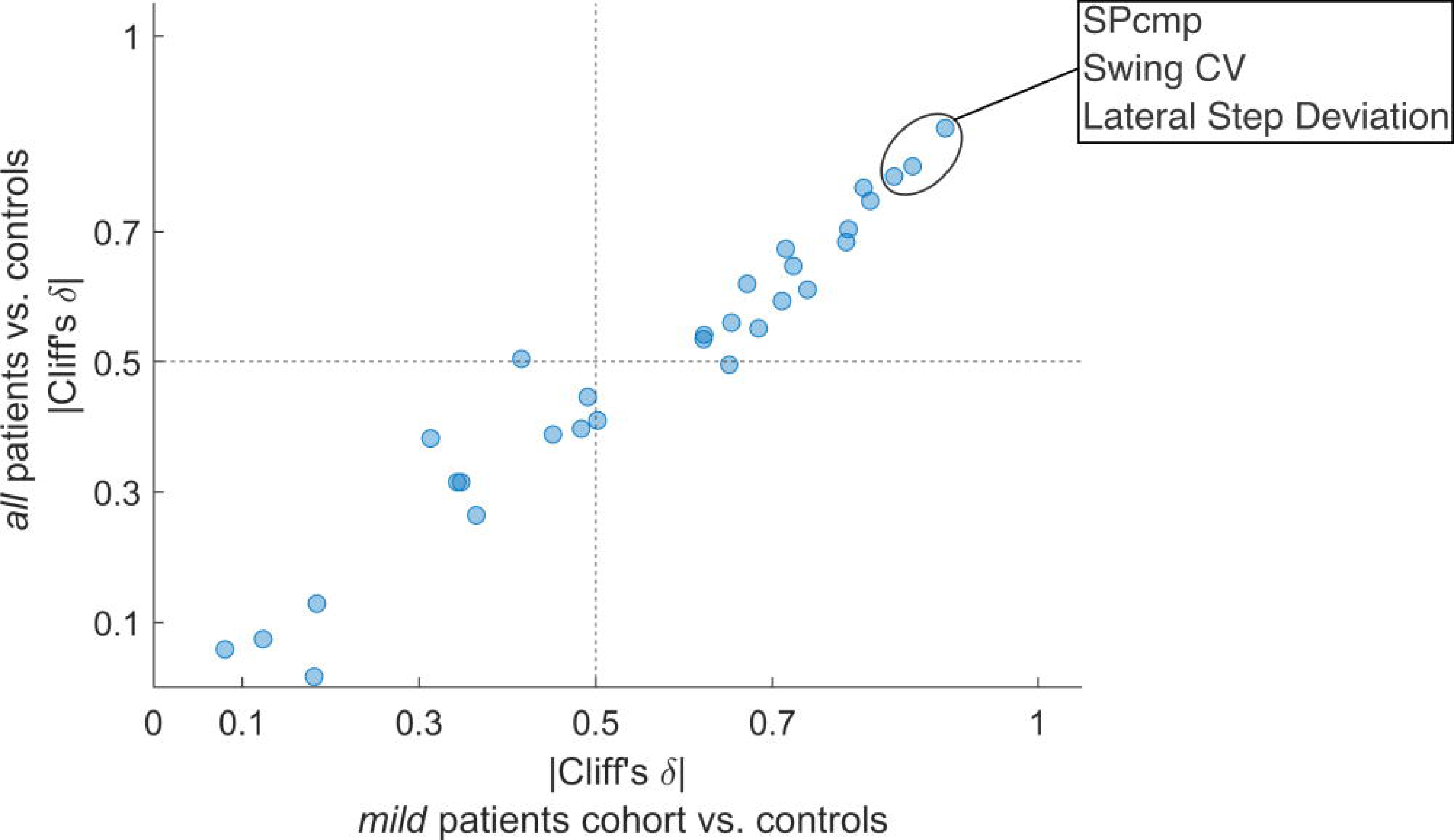
Discriminatory power of 30 gait measures in lab-based walking. Scatter plot displaying discriminative effects size Cliff’s δ for all SPG7 patients (y-axis) and the mild patient cohort (SPRS^mobility^ ≤ 9, x-axis) against healthy controls for each gait measure. Top 3 measures are highlighted.

**Table 1:** Discrimination between SPG7 patients and HC (lab-based walking)

|  | SPG7 |  | HC |  |  |  |
| --- | --- | --- | --- | --- | --- | --- |
| *N (f/m)* | 65 (20/45) |  | 50 (27/23) |  |  |  |
| ***Demographic/clinical measures*** | Median | MADN | Median | MADN |  |  |
| Age (y) | 53.0 | 10.4 | 47.5 | 17.8 |  |  |
| Disease duration (y) | 15.0 | 7.4 |  |  |  |  |
| SPRS<sup>mobility</sup> | 9.00 | 2.97 | 0.00 | 0.00 |  |  |
| SPRS | 14.0 | 4.4 | 0.00 | 0.00 |  |  |
| SARA<sup>PG</sup> | 4.00 | 1.48 | 0.00 | 0.00 |  |  |
| SARA | 9.00 | 3.71 | 0.00 | 0.00 |  |  |
| FARS-ADL | 9.00 | 4.45 | 0.00 | 0.00 |  |  |
| ***Gait measures*** | Median | MADN | Median | MADN | Cliff's $\delta$ | p-value |
| SPcmp | 0.361 | 0.146 | 0.158 | 0.066 | 0.90 | 2.3e-16 * |
| Swing CV | 0.0236 | 0.0109 | 0.0121 | 0.0034 | 0.86 | 3.6e-15 * |
| Lateral Step Deviation (%) | 3.48 | 1.18 | 1.94 | 0.52 | 0.84 | 1.6e-14 * |
| Pitch at Initial Contact (°) | 18.7 | 5.7 | 27.5 | 4.8 | -0.81 | 7.4e-13 * |
| Harmonic Ratio V | 2.02 | 0.52 | 3.63 | 0.84 | -0.80 | 1.8e-13 * |
| Harmonic Ratio AP | 1.90 | 0.48 | 3.19 | 0.91 | -0.79 | 5.9e-13 * |
| Pitch at Toe Off (°) | 29.5 | 6.8 | 37.9 | 4.0 | -0.78 | 7e-13 * |
| Stride Time CV | 0.0329 | 0.0176 | 0.0169 | 0.0067 | 0.74 | 1.2e-11 * |
| Circumduction | 5.05 | 1.83 | 2.79 | 0.95 | 0.72 | 3.3e-11 * |
| Double Support MADN | 1.48 | 0.53 | 0.887 | 0.219 | 0.72 | 5.6e-11 * |
| Stride Length (m) | 1.05 | 0.20 | 1.33 | 0.19 | -0.71 | 7.3e-11 * |
| Gait Speed (m/s) | 0.924 | 0.219 | 1.24 | 0.20 | -0.68 | 3.6e-10 * |
| Harmonic Ratio ML | 1.66 | 0.56 | 2.47 | 0.64 | -0.67 | 7.6e-10 * |
| Pitch at Toe Off MADN | 1.35 | 0.48 | 0.791 | 0.295 | 0.65 | 2.1e-09 * |
| Stride Length CV | 0.0423 | 0.0182 | 0.0255 | 0.0074 | 0.65 | 2.4e-09 * |
| Double Support (%) | 25.4 | 4.7 | 19.5 | 2.7 | 0.62 | 1.2e-08 * |
| Swing (%) | 37.4 | 2.4 | 40.3 | 1.4 | -0.62 | 1.2e-08 * |
| Pitch at Initial Contact MADN | 1.55 | 0.57 | 1.05 | 0.36 | 0.50 | 9e-06 * |
| LROM transverse (°) | 12.7 | 5.4 | 8.94 | 2.70 | 0.49 | 6.8e-06 * |
| Toe Out Angle MADN | 2.11 | 0.90 | 1.47 | 0.66 | 0.48 | 9.4e-06 * |
| Elevation at Midswing MADN | 0.358 | 0.145 | 0.235 | 0.067 | 0.45 | 3.5e-05 * |
| Pitch at Mid Swing (°) | 11.3 | 4.6 | 15.1 | 3.8 | -0.42 | 0.00014 * |
| Stride Time (s) | 1.15 | 0.11 | 1.07 | 0.06 | 0.36 | 0.00083 * |
| LROM sagittal (°) | 6.58 | 2.36 | 4.78 | 1.42 | 0.35 | 0.0014 * |
| LROM sagittal CV | 0.180 | 0.063 | 0.138 | 0.042 | 0.34 | 0.0017 * |
| Elevation at Midswing (cm) | 1.57 | 0.73 | 1.18 | 0.48 | 0.31 | 0.0041 |
| Toe Out Angle (°) | 9.86 | 5.18 | 8.02 | 5.53 | 0.18 | 0.091 |
| LROM coronal CV | 0.0952 | 0.0508 | 0.0896 | 0.0401 | 0.18 | 0.097 |
| LROM coronal (°) | 7.20 | 2.12 | 7.69 | 2.18 | -0.12 | 0.26 |
| LROM transverse CV | 0.192 | 0.071 | 0.184 | 0.071 | 0.08 | 0.46 |

\* $p < 0.05/n=30$ : # of gait measures (Bonferroni). HC: healthy controls. SPRS: Spastic Paraplegia Rating Scale. SPRS<sup>mobility</sup>: mobility subscore of the SPRS (items 1-6). SARA: Scale for the Assessment and Rating of Ataxia. SARA<sup>PG</sup>: posture&gait subscore of SARA (items 1-4). FARS-ADL: activities of daily living subscore of the Friedreich Ataxia Rating Scale. MADN: normalized median absolute deviation.

To evaluate the ability of the candidate measures to discriminate patients with mild disease severity from HC, we performed a median split of the patient cohort with respect to the SPRS^mobility^ scale. For this mild patient cohort (SPRS^mobility^ ≤ 9, n=41) discrimination with large effect sizes was observed for 2/30 measures, and at least moderate effect sizes for 17/30 measures (supplementary table 3). The strongest discrimination was observed for the measures SPcmp (δ = 0.86), Swing CV (δ = 0.80), and Lateral Step Deviation (δ = 0.78), mirroring the results for the whole patient cohort with minor decreases in effect size (figure 1). Comparing the sets of measures with at least moderate effect size between the whole patient cohort and the mild patient cohort, a large overlap was apparent (16 measures identical). We did not identify a measure that exhibited large discriminative power only in mild patient cohort.

### Measures of spatiotemporal gait variability correlate with clinical measures of mobility even in patients with mild disease severity

For the at least moderately discriminative gait measures in the whole patient cohort (18 measures) and mild patient cohort (17 measures), we assessed correlations with clinician-reported measures (primary outomes: SPRS^mobility^, SARA^PG^; exploratory outcomes: SPRS, SARA, FARS-ADL). Discriminative gait measures correlated with clinician-reported measures of mobility, and posture and gait, with large effect sizes. For the mobility scale SPRS^mobility^, the largest effect sizes were observed for correlations with measures of spatiotemporal gait variability such as the spatial variability composite measure SPcmp (ρ = 0.67, p = 9.1e-10), Stride Time CV (ρ = 0.67, p = 1.5e-9) and Swing CV (ρ = 0.64, p = 1.1e-8). (figure 2 and table 2) The same gait measures also correlated with SARA^PG^, with only minor differences of effect sizes. For the gait measures Stride Length, Gait Speed, Pitch at Toe Off, Double Support and Swing, however, significantly larger effect sizes were observed for the correlations with SPRS^mobility^ than with SARA^PG^. On the other hand, the measure of foot angle variability Pitch at Toe Off MADN correlated more strongly with SARA^PG^ than with SPRS^mobility^.

**Figure 2:**
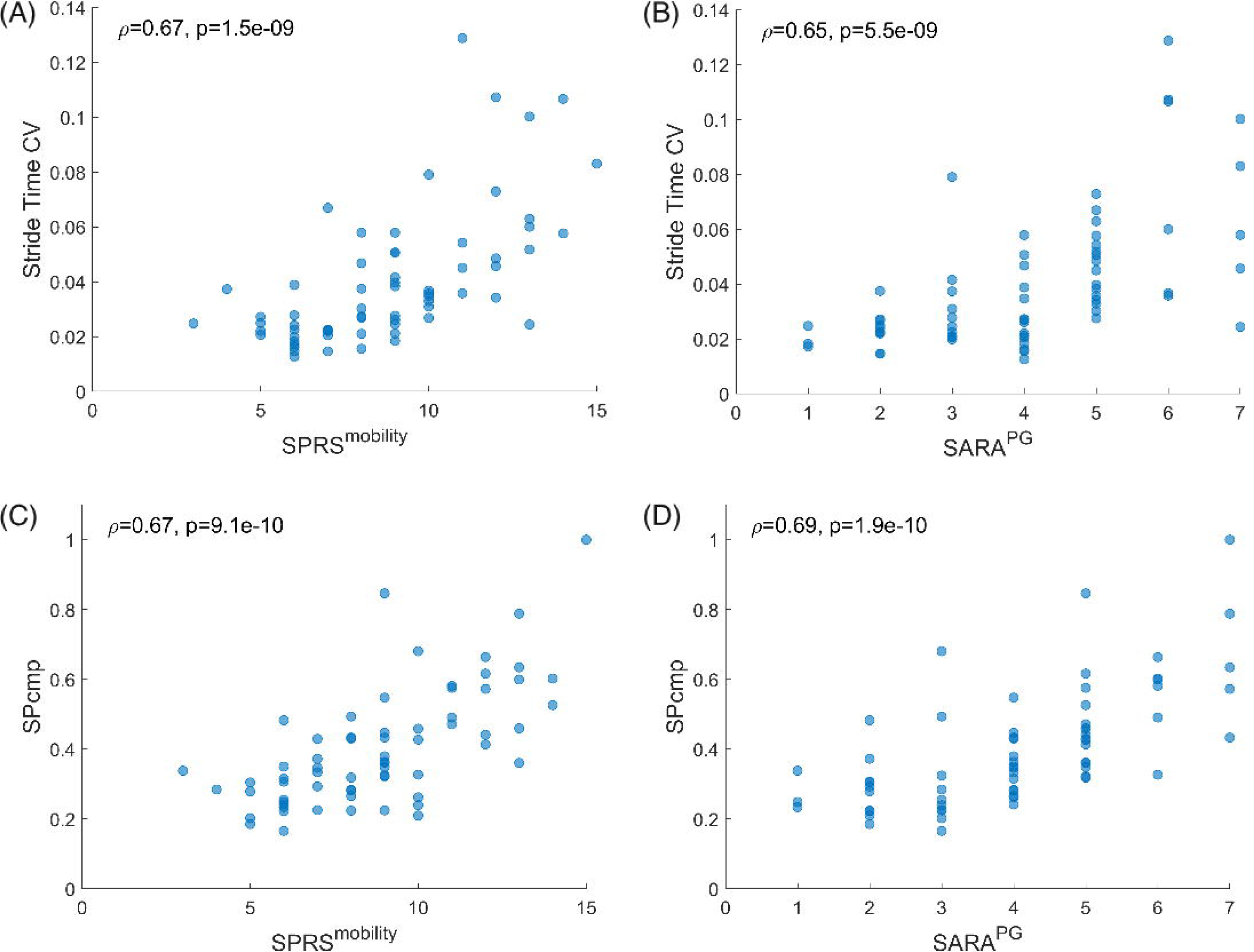
Top correlations between gait measures and clinical measures in all SPG7 patients during lab- based walking: (A, B) Stride Time CV and (C, D) SPcmp vs. SPRS^mobility^ (left) and SARA^PG^ (right). Spearman’s ρ and associated p value depicted in upper left corner.

**Table 2:**
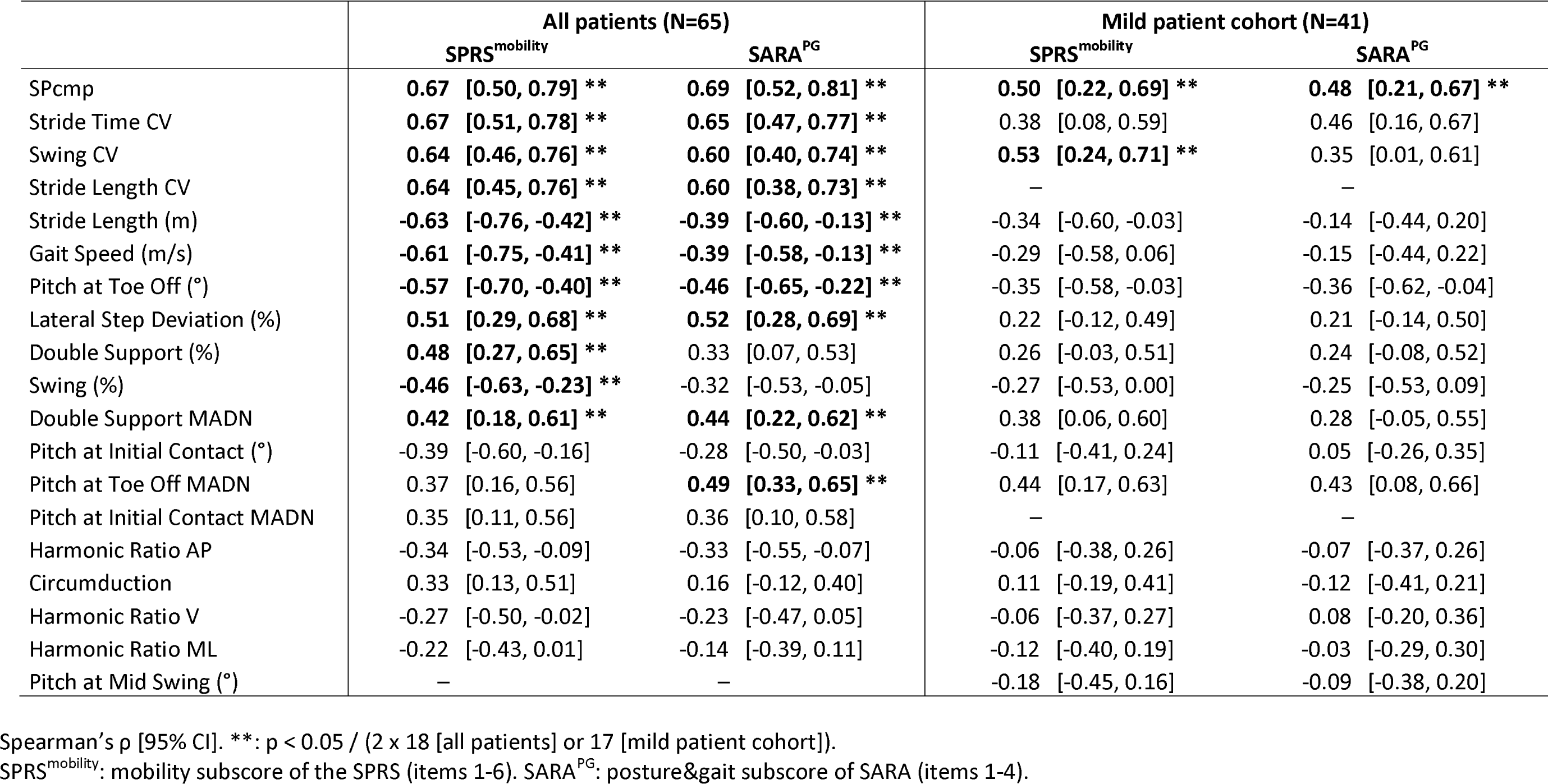
Correlation of discriminative gait measures with primary clinical measures (lab-based walking; all patients [left] and mild patient cohort [right])

|  | All patients (N=65) |  | Mild patient cohort (N=41) |  |
| --- | --- | --- | --- | --- |
|  | SPRS<sup>mobility</sup> | SARA<sup>PG</sup> | SPRS<sup>mobility</sup> | SARA<sup>PG</sup> |
| SPcmp | **0.67** [0.50, 0.79] ** | **0.69** [0.52, 0.81] ** | **0.50** [0.22, 0.69] ** | **0.48** [0.21, 0.67] ** |
| Stride Time CV | **0.67** [0.51, 0.78] ** | **0.65** [0.47, 0.77] ** | 0.38 [0.08, 0.59] | 0.46 [0.16, 0.67] |
| Swing CV | **0.64** [0.46, 0.76] ** | **0.60** [0.40, 0.74] ** | **0.53** [0.24, 0.71] ** | 0.35 [0.01, 0.61] |
| Stride Length CV | **0.64** [0.45, 0.76] ** | **0.60** [0.38, 0.73] ** | – | – |
| Stride Length (m) | **-0.63** [-0.76, -0.42] ** | **-0.39** [-0.60, -0.13] ** | -0.34 [-0.60, -0.03] | -0.14 [-0.44, 0.20] |
| Gait Speed (m/s) | **-0.61** [-0.75, -0.41] ** | **-0.39** [-0.58, -0.13] ** | -0.29 [-0.58, 0.06] | -0.15 [-0.44, 0.22] |
| Pitch at Toe Off (°) | **-0.57** [-0.70, -0.40] ** | **-0.46** [-0.65, -0.22] ** | -0.35 [-0.58, -0.03] | -0.36 [-0.62, -0.04] |
| Lateral Step Deviation (%) | **0.51** [0.29, 0.68] ** | **0.52** [0.28, 0.69] ** | 0.22 [-0.12, 0.49] | 0.21 [-0.14, 0.50] |
| Double Support (%) | **0.48** [0.27, 0.65] ** | 0.33 [0.07, 0.53] | 0.26 [-0.03, 0.51] | 0.24 [-0.08, 0.52] |
| Swing (%) | **-0.46** [-0.63, -0.23] ** | -0.32 [-0.53, -0.05] | -0.27 [-0.53, 0.00] | -0.25 [-0.53, 0.09] |
| Double Support MADN | **0.42** [0.18, 0.61] ** | **0.44** [0.22, 0.62] ** | 0.38 [0.06, 0.60] | 0.28 [-0.05, 0.55] |
| Pitch at Initial Contact (°) | -0.39 [-0.60, -0.16] | -0.28 [-0.50, -0.03] | -0.11 [-0.41, 0.24] | 0.05 [-0.26, 0.35] |
| Pitch at Toe Off MADN | 0.37 [0.16, 0.56] | **0.49** [0.33, 0.65] ** | 0.44 [0.17, 0.63] | 0.43 [0.08, 0.66] |
| Pitch at Initial Contact MADN | 0.35 [0.11, 0.56] | 0.36 [0.10, 0.58] | – | – |
| Harmonic Ratio AP | -0.34 [-0.53, -0.09] | -0.33 [-0.55, -0.07] | -0.06 [-0.38, 0.26] | -0.07 [-0.37, 0.26] |
| Circumduction | 0.33 [0.13, 0.51] | 0.16 [-0.12, 0.40] | 0.11 [-0.19, 0.41] | -0.12 [-0.41, 0.21] |
| Harmonic Ratio V | -0.27 [-0.50, -0.02] | -0.23 [-0.47, 0.05] | -0.06 [-0.37, 0.27] | 0.08 [-0.20, 0.36] |
| Harmonic Ratio ML | -0.22 [-0.43, 0.01] | -0.14 [-0.39, 0.11] | -0.12 [-0.40, 0.19] | -0.03 [-0.29, 0.30] |
| Pitch at Mid Swing (°) | – | – | -0.18 [-0.45, 0.16] | -0.09 [-0.38, 0.20] |

Spearman's $\rho$ [95% CI]. \*\*: $p < 0.05$ / (2 x 18 [all patients] or 17 [mild patient cohort]).
SPRS<sup>mobility</sup>: mobility subscore of the SPRS (items 1-6). SARA<sup>PG</sup>: posture&gait subscore of SARA (items 1-4).

Even within the mild patient cohort, correlations between gait measures and clinician-reported measures of mobility, posture and gait were observed with medium to large effect sizes. Specifically, three gait measures correlated with SPRS^mobility^ – Swing CV (ρ = 0.53, p = 4.2e-4), SPcmp (ρ = 0.50, p = 9.5e-4), and Stride Length CV (ρ = 0.48, p = 0.0014) (for visualisation see supplementary figure 3) – and one gait measure correlated with SARA^PG^ – SPcmp (ρ = 0.48, p = 0.0014) (table 2). Of note, within the mild patient cohort all gait measures with significant correlations belonged to the domain of spatiotemporal gait variability.

For standard clinical measures of HSP-related and ataxia-related disease severity, correlations with large effect sizes of ρ ≥ 0.5 were primarily observed for measures of spatiotemporal gait variability (supplementary table 4). Specifically, the correlations of the largest effect sizes with HSP-related disease severity (SPRS) were observed for SPcmp, Stride Time CV and Stride Length CV. The measures Stride Length CV, Stride Time CV and Swing CV displayed the largest effect sizes in the correlation with ataxia-related disease severity (SARA). Several of the measures correlated with a measure of patient-relevant impairment in everyday life activities (FARS-ADL), with Stride Length CV (ρ = 0.45, p = 1.9e-4), Swing CV (ρ = 0.44, p = 2.7e-4) and Gait Speed (ρ = -0.41, p = 7.6e-4) leading the list. Also the previously highlighted measures Stride Time CV (ρ = 0.38, p = 0.0019) and SPcmp (ρ = 0.39, p = 0.0015) correlated with FARS-ADL (supplementary figure 4).

Comparison of groups of patients in different disease stages defined by the FARS Staging (mild: ≤ 2.0, intermediate: 2.5-3.5, advanced: ≥ 4.0) revealed significant group differences between mild, intermediate, and advanced stages for SPcmp (p_Kruskal-Wallis_=1.1e-4, p_mild-intermediate_=7.4e-4, p_intermediate-advanced_=0.022) and between mild and intermediate stages for Stride Time CV (p_Kruskal-Wallis_=2.2e-4, p_mild-intermediate_=8.6e-5, p_intermediate-advanced_=0.71). Figure 3 illustrates the thus defined approximate meaningful score regions of the two gait measures.

**Figure 3:**
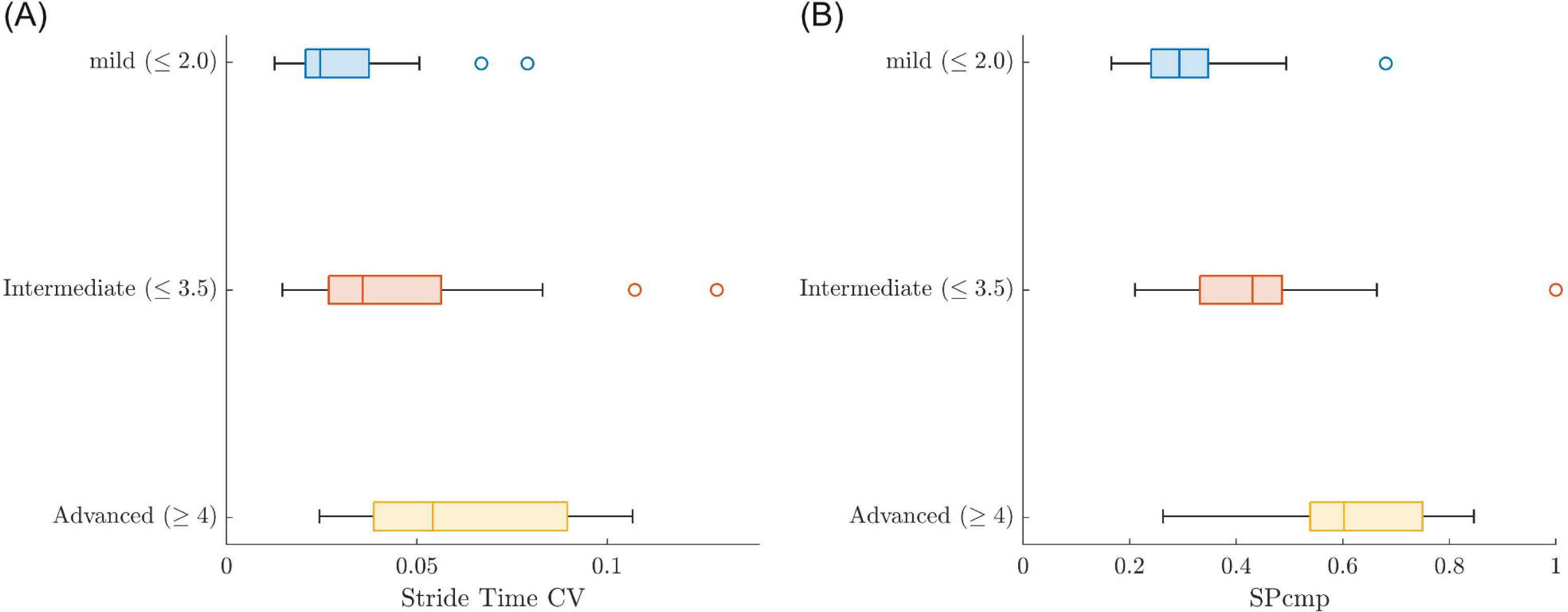
Box plots with values of (A) Stride Time CV and (B) SPcmp in groups of patients in different disease stages, defined by FARS Staging (mild: ≤2; intermediate: 2.5-3.5; advanced: ≥4). Line inside Boxes = median, edges of boxes = lower & upper quartile, whiskers = (non-outlier) minimum & maximum, circles = outliers (values > 1.5×interquartile range away from lower/upper edge of box).

### Measures of pace and gait variability discriminate patients from controls and correlate with clinician-reported measures of mobility under conditions simulating real life

To assess performance of gait measures in simulated real life, discrimination of patients from controls and correlation with clinician-reported outcomes were assessed for data recorded under the SFW condition. Gait measures discriminated patients from HC in SFW with large effect sizes for 3/30 measures and with at least moderate effect sizes for 17/30 measures (table 3). The most discriminative measures – Harmonic Ratio V (δ = -0.85), Harmonic Ratio AP (δ = -0.80), Swing CV (δ = 0.83) – capture gait smoothness and temporal variability. In contrast to LBW, measures of spatial gait variability (Lateral Step Deviation, Stride Length CV, SPcmp) were not among the top measures but still discriminated with moderate effect sizes.

**Table 3:**
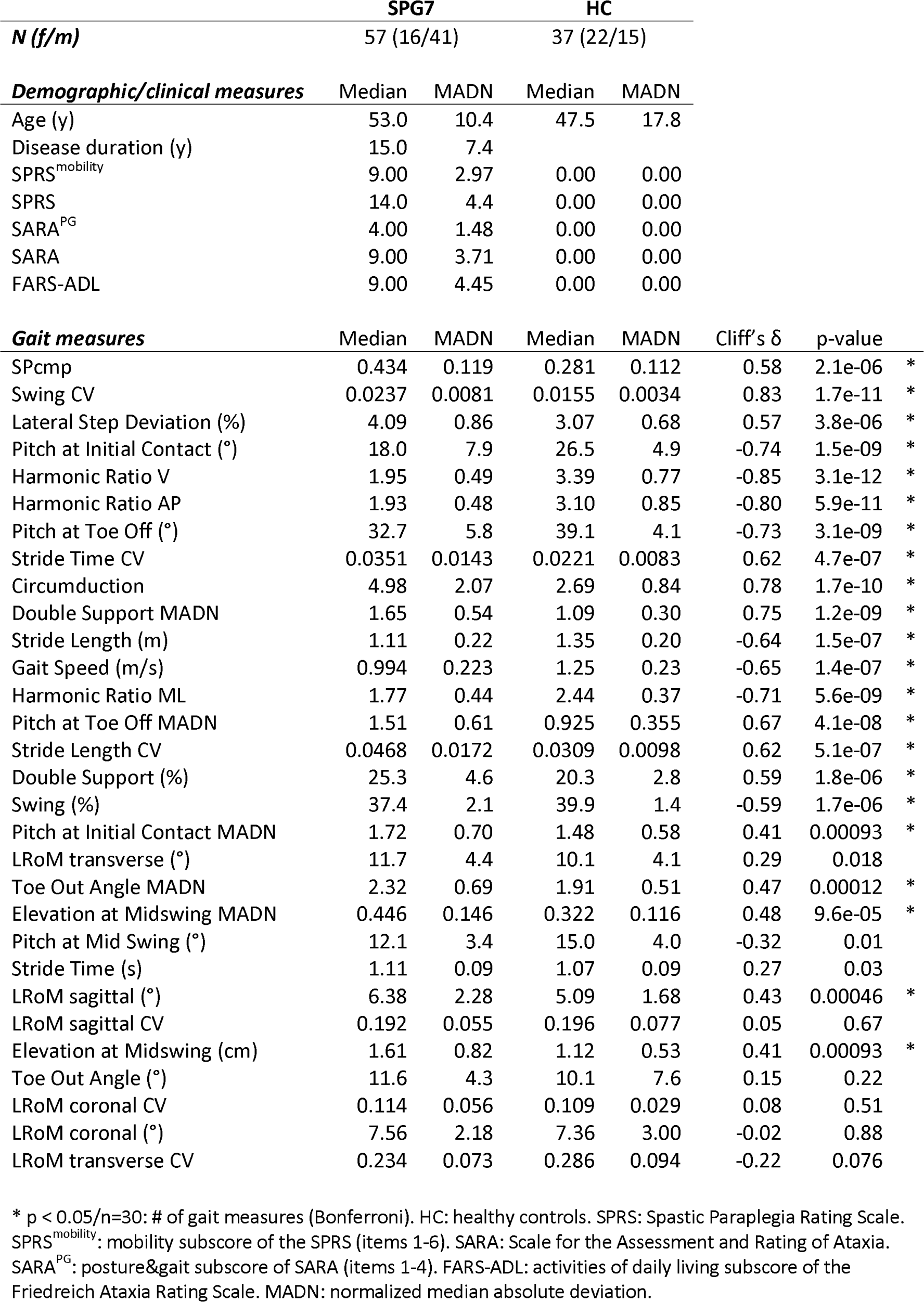
Discrimination of SPG7 patients from HC (supervised free walking)

At least large correlations between the LBW and SFW conditions were observed for 27/30 gait measures (very large: 17, large: 10, medium: 3; supplementary table 6). Consequently, largely the same set of measures discriminated patients from controls in SFW as in LBW (though the rankings within the set of discriminative measures differed between the two conditions). Specifically, except Pitch at Initial Contact MADN, all measures discriminative with at least moderate effect size in LBW were reproduced in SFW. Reversely, all measures with at least moderate effect sizes in SFW were also found to be discriminative in LBW.

Gait measures discriminative in SFW correlated with the mobility measure SPRS^mobility^ with large effect sizes. In contrast to the results for LBW, two out of the three top measures – Gait Speed (ρ = -0.59, p = 1.1e-6), Stride Time CV (ρ = 0.57, p = 3.7e-06) and Stride Length (ρ = -0.55, p = 1.2e-05) – were measures of pace rather than gait variability (figure 4, supplementary table 7). Still, all measures of spatiotemporal gait variability correlated with SPRS^mobility^, except for Lateral Step Deviation. Correlations with SARA^PG^ were found for the measures Stride Length CV (ρ = 0.47, p = 1.9e-04), Double Support MADN (ρ = 0.46, p = 3.0e-4) and Pitch at Toe Off MADN (ρ = 0.46, p = 3.2e-4).

**Figure 4:**
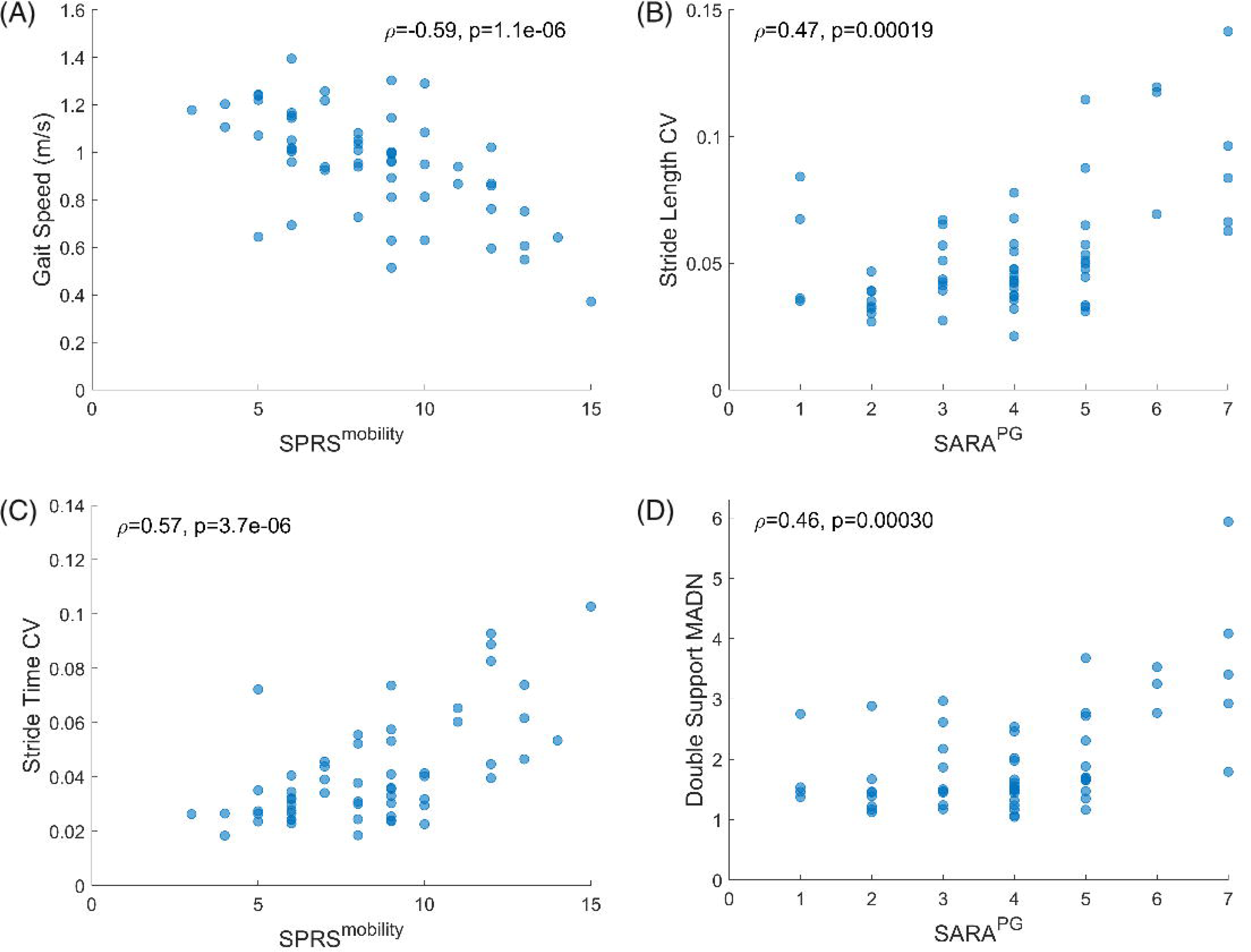
Top correlations between gait measures and clinical measures in all SPG7 patients during supervised free walking: (A) Gait Speed and (C) Stride Time CV vs. SPRS^mobility^; (B) Stride Length CV and (D) Double Support MADN vs. SARA^PG^. Spearman’s ρ and associated p value depicted in upper right/left corner.

Top measures in the correlation with HSP-related disease severity (SPRS) were Stride Time CV, Gait Speed and Pitch at Initial Contact, all displaying moderate effect sizes (supplementary table 7). Correlations with ataxia-related disease severity (SARA) were found for Double Support MADN, Pitch at Toe Off MADN – both with large effect sizes – and Stride Length CV. Correlations with FARS-ADL were found for measures of pace and temporal and foot angle variability, with moderate effect sizes for Gait Speed and Double Support.

## Discussion

In this cross-sectional, multi-center study, we employed body-worn sensors to identify candidate digital gait outcomes for upcoming treatment trials for SPG7. The study’s design, encompassing data capture across multiple sites, and gait assessments both in laboratory and simulated real-life conditions, closely mirrored anticipated settings in future clinical trials. We established a shortlist of promising gait measures through a structured multistep approach. Beginning with a hypothesis-based pre-selection of gait measures informed by the literature and clinical experience, we subsequently narrowed down the list to measures with high discriminative power, finally selecting those that exhibited the strongest correlation with clinical measures of patient-relevant health aspects.

The gait measures that demonstrated the largest correlations with the SPRS^mobility^ and SARA^PG^ were related to spatiotemporal gait variability, as exemplified by the spatial variability composite measure SPcmp (SPRS^mobility^: Spearman’s ρ = 0.67; SARA^PG^: ρ = 0.69) and Stride Time CV (SPRS^mobility^: ρ = 0.67; SARA^PG^: ρ = 0.65). At the same time, these parameters also discriminated between patients and controls (SPcmp: Cliff’s δ = 0.90; Stride Time CV: δ = 0.74). These results corroborate and extend findings from previous studies performed in mixed or genotype-specific cohorts of patients with cerebellar ataxias. In a previous study with a mixed cohort of patients with cerebellar ataxias, gait measures such as variability of stride length and of stride duration have demonstrated discriminative power between patients and controls, and have shown correlations with the SARA^PG^ (Ilg et al., 2020). In another study with a mixed cohort of SCA patients, spatial and temporal stride variability measures discriminated between patients and controls (Shah et al., 2021). In the same study, variability of the double support phase was among the measures that correlated most strongly with the SARA in SCA patients. The other top measures were, however, related to foot angles and their variability, measures that were not among the prime candidates in the present SPG7 study.

Our results, highlighting measures of spatiotemporal variability as prime candidate gait outcomes, also extend the findings of the very few published studies employing sensor-based gait analysis in HSP. In the largest study, measures of temporal stride variability such as Stride Time CV discriminated patients from controls and demonstrated cross-sectional correlation with SPRS (Regensburger et al., 2022). However, they did not show longitudinal progression on follow-up assessments (Loris et al., 2023). The cited studies were limited, however, by the inclusion of a genotypically mixed and thus phenotypically heterogeneous cohort of HSP patients. The cohort included 17 SPG7 patients, but no genotype-specific subgroup analysis was presented. The study was further limited by analyzing only temporal and gait cycle measures, and not spatial or foot angle measures. Moreover, it included patients dependent on walking aids – also during the measurements, which may have a profound impact on gait measures – and was thus of reduced informative value for trial-relevant mild disease stages.

Taken together, our results extend the utility of spatiotemporal gait variability measures to capture disease-related gait impairment from cerebellar ataxias – and from HSP, though here only partially and much less established – to the spastic ataxia SPG7. *A* priori, this transferability was not self-evident, as it was unclear how the variable spastic component of gait in SPG7 adds to or modifies gait features one would observe in pure ataxic gait and vice versa. This finding may extend to other spastic ataxias, making the gait measures identified in this study potential candidate outcomes for other genotypes as well.

Several gait measures evaluated in this study discriminated between SPG7 patients with mild disease severity and controls and demonstrated correlations with clinician-reported outcome assessments even *within* this mild cohort. This is of particular importance as future disease-modifying treatment trials will likely focus on patients in mild disease stages. Widely used clinical outcome scales in ataxia and HSP – SARA and SPRS – have been designed to capture the full disease spectrum. Therefore, they may have limited sensitivity to change and are prone to floor effects in these early stages. Digital gait measures on the other hand may allow monitoring of disease progression within trial-relevant time frames of one to two years even within patients with mild disease (Seemann et al., 2023). The gait measures identified in this study, demonstrating strong discriminative power and correlation with clinical scales in mild disease stages, are thus prime candidate outcomes for potential treatment trials.

To serve as meaningful outcomes in treatment trials, digital-motor measures must not merely capture change over time but also reflect changes relevant to patients. Mobility is a concept that is known to be of particularly high relevance for patients with HSP (Malina et al., in press) as well as ataxia (Gorcenco et al., 2022; Lowit et al., 2021; Trace et al., 2021). The robust correlations with the SPRS^mobility^ scale therefore indicate that our top gait measures indeed mirror disease aspects that are meaningful to patients. However, the extent to which gait assessments in controlled laboratory conditions accurately capture aspects of mobility that are relevant to patients in real life was unknown. To address this limitation, this study included assessments of ‘supervised free walking’ conducted outside the laboratory but within institutional compounds and under supervision by staff members, thus simulating more complex real-life conditions while concurrently maintaining stringent control to ensure technical robustness. Remarkably, gait measures discriminated patients and controls and correlated with clinical measures in the SFW condition even though the walking routes naturally differed between the multiple participating centers. The successful application of this assessment approach in SPG7 opens up two distinct perspectives: Firstly, by showing that relevant gait measures could be captured in more variable conditions simulating real life, this study paves the way towards real-life measurements in patients’ everyday lives, which in turn would offer a maximum of ecological validity. Secondly, assessments of ‘supervised free walking’ could serve as a means to increase ecological validity of digital gait outcomes in multi-center treatment trials while simultaneously avoiding the technical challenges associated with real-life measurements.

The two top gait measures, Stride Time CV and SPcmp, differentiated between groups of patients defined by a staging of disease severity on a functional, patient-relevant level (FARS Staging). The ranges of values associated with each disease stage could thus be interpreted as approximate meaningful score regions, a concept that has recently been endorsed by the FDA ((FDA), 2023). Consequently, establishing the association between ranges of values of gait measures and disease stages defined through patient-relevant health aspects underlines the patient-meaningfulness of these gait measures.

This study has several limitations. Due to its cross-sectional design, it could not evaluate responsiveness to change, a crucial criterion for the viability of gait measures as outcomes in treatment trials. Consequently, the measures identified here will need to be evaluated longitudinally. Furthermore, while the results of this study indicate that the identified gait measures could reflect health aspects that are meaningful to patients, more work is needed to establish the meaningfulness of these gait measures.

In conclusion, this study identified multi-center applicable digital gait measures capable of discriminating between patients and controls and correlating with relevant COAs, even in mild disease stages, and in settings simulating real-life. If validated longitudinally, the measures are prime candidate outcomes for future treatment trials for SPG7 and other spastic ataxias.

## Data Availability

Data will be made available upon reasonable request and as patient consent allows.

## Acknowledgement

This work was supported by the DFG under the frame of EJP-RD network PROSPAX (No 441409627; to R.S., M.S., IR, FMS) and by the Clinician Scientist program “PRECISE.net” funded by the Else Kröner-Fresenius-Stiftung (to L.B., A.T., and M.S.). The study was further funded by the German Ministry for Education and Research (BMBF) via funding for the TreatHSP network (01GM1905 to R.S. and S.K.; 01GM2209A to R.S.). J.S. was supported by the International Max Planck Research School for Intelligent Systems (IMPRS-IS) and the Else Kröner-Fresenius-Stiftung Medical Scientist programme ‘ClinbrAIn’. A.T. received funding from the University of Tübingen, medical faculty, for the Clinician Scientist Program Grant #439–0–0. I.R. is supported in part by the Italian Ministry of Health (Ricerca Finalizzata RF-2019-12370417; Ricerca Corrente 2023, RC 5x1000). A.N.B. is supported by TUBITAK (grant no: 319S063). B.vdW. is supported by ZonMw (the Netherlands Organization for Scientific Research; 463002002). The movement analysis data from İstanbul site used in this study was conducted using the service and infrastructure of Koç University Research Center for Translational Medicine (KUTTAM). We are grateful to Selina Reich for her coordinating support as part of the PROSPAX consortium and to Tanja Heger for monitoring the datasets of the PROSPAX registry. We would like to thank Ilse Willemse for conducting sensor measurements within the Nijmegen cohort. Several authors are part of the European Reference Network for Rare Neurological Diseases (ERN-RND; M.S., B.vdW., R.S.).

## Disclosure

L.B., J.S., C.K., A.T., D.M., K.D.-J., I.R., S.S., A.N.B., G.C., D.T., C.G., R.S. report no disclosures relevant to the manuscript. B.vdW. receives research support from ZonMw, The Netherlands Organization for Scientific Research, Hersenstichting, Christina Foundation, and Radboud university medical center; B.vdW. has served on scientific advisory boards or provided consultancy services to Vico Therapeutics, Servier, and Biohaven Pharmaceuticals; B.vdW. has received royalties from BSL-Springer Nature and speaker fees from MDC Malaysia. W.I. received consultancy honoraria from Ionis Pharmaceuticals. M.S. has received consultancy honoraria from Ionis, UCB, Prevail, Orphazyme, Servier, Reata, GenOrph, AviadoBio, Biohaven, Zevra, and Lilly, all unrelated to the present manuscript.

## Author roles

1. Research project: A. Conception, B. Organization, C. Execution;

2. Statistical Analysis: A. Design, B. Execution, C. Review and Critique;

3. Manuscript Preparation: A. Writing of the first draft, B. Review and Critique;

L.B.: 2A, 2B, 3A

J.S.: 2C, 3B

C.K.: 1C, 3B

A.T.: 1C, 3B

D.M.: 1B

K.D.-J.: 1B

I.R.: 1C, 3B

S.S.: 1C, 3B

A.N.B.: 1C, 3B

G.C.: 1C, 3B

D.T.: 1C, 3B

C.G.: 1C, 3B

B.vdW.: 1C, 3B

W.I.: 2A, 2C, 3B

M.S.: 1A, 1B, 2A, 2C, 3A, 3B

R.S.: 1A, 1B, 2A, 2C, 3A, 3B

## Additional study group contributors of the PROSPAX consortium

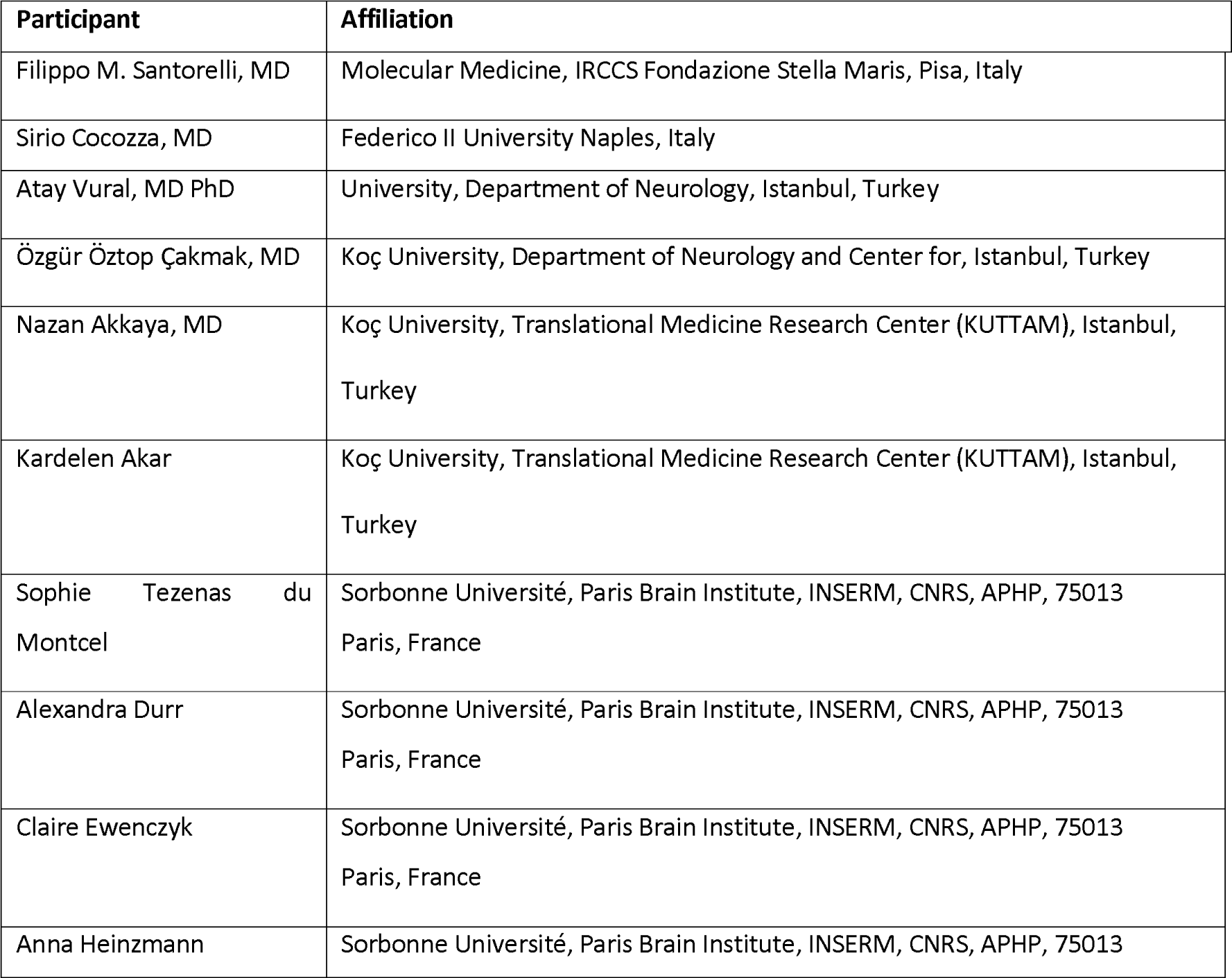
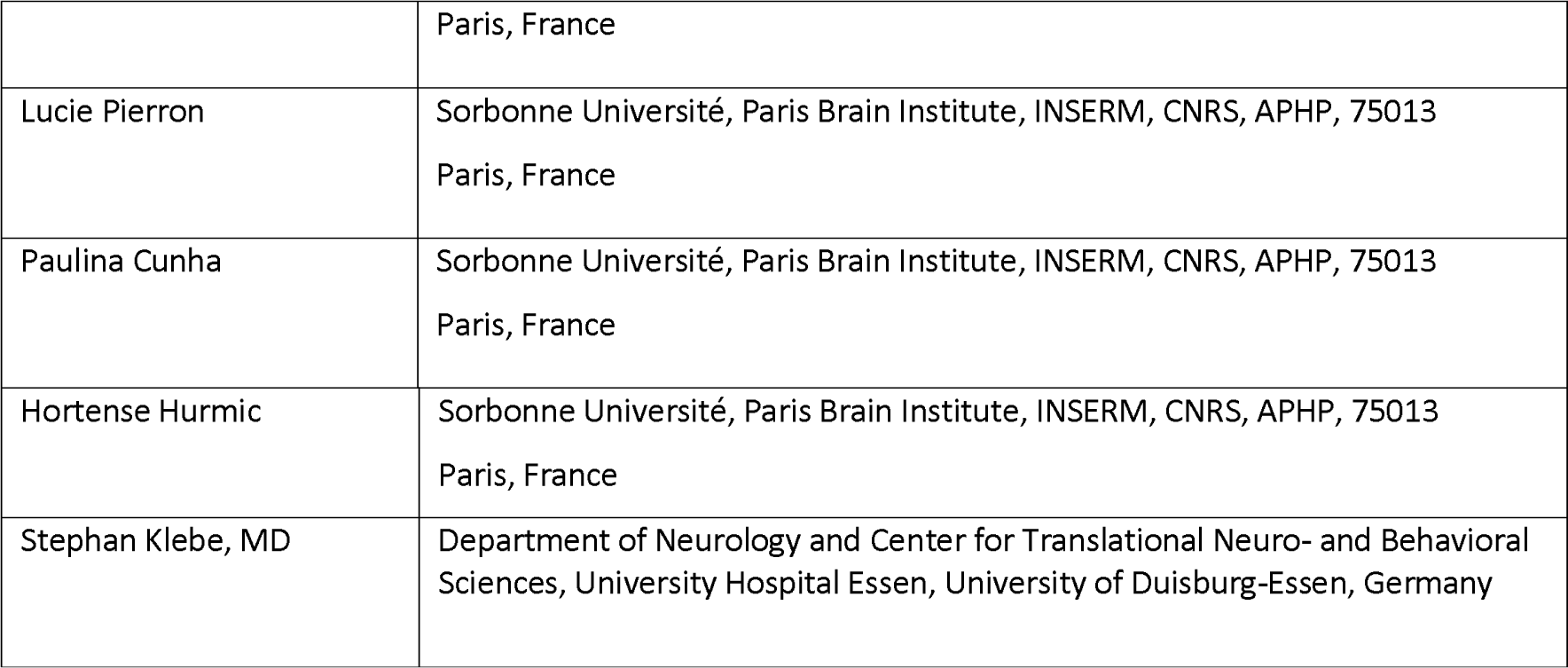

## Notes

### Competing Interest Statement

The authors have declared no competing interest.

### Author Declarations

The Institutional Review Board of the University of Tuebingen gave ethical approval for this work (AZ 824/2019BO2)

