## supplementary_tables for "Towards patient-relevant, trial-ready digital motor outcomes for SPG7: a cross-sectional prospective multi-center study (PROSPAX)"

**Supplementary table 1: Gait measures analyzed and their definitions**

| **Gait measures** | **Definition** | **Measure included in analysis** | | |
| --- | --- | --- | --- | --- |
|  |  | **Median** | **MADN** | **CV** |
| Stride Time (s) | The duration of a full gait cycle, measured from the left foot’s initial contact to the next initial contact of the left foot |  |  | **×** |
| Stride Length (m) | The forward distance travelled by a foot during a gait cycle | **×** |  | **×** |
| Gait Speed (m/s) | The forward speed of the subject, measured as the forward distance travelled during gait cycle divided by gait cycle duration | **×** |  |  |
| Lateral Step Deviation  (% of Stride Length) | In a series of 3 consecutive foot placements of the same foot, the perpendicular deviations of the middle foot placement from the line connecting the ﬁrst and third, normalized by stride length | **×** |  |  |
| Pitch at Toe Off (°) | The angle of the foot as it leaves the ﬂoor at push-off. The pitch of the foot when ﬂat is zero. | **×** | **×** |  |
| Pitch at Midswing (°) | The angle of the foot at midswing. | **×** |  |  |
| Pitch at Initial Contact (°) | The angle of the foot at the point of initial contact. The pitch of the foot when ﬂat is zero and positive when the heel contacts ﬁrst. | **×** | **×** |  |
| Toe Out Angle (°) | The lateral angle of the foot during the stance phase, relative to the forward motion of the gait cycle. Positive angle is outward rotation. | **×** | **×** |  |
| Elevation at Midswing (cm) | The height of the foot sensor measured at midswing, relative to its start position while standing | **×** | **×** |  |
| Circumduction  (% of stride length) | The amount that the foot travels perpendicular to forward movement while swinging forward during an individual stride, normalized by stride length | **×** |  |  |
| Double Support  (% of gait cycle) | The percentage of the gait cycle in which both feet are on the ground | **×** | **×** |  |
| Swing (% of gait cycle) | The percentage of the gait cycle in which the foot is not on the ground | **×** |  | **×** |
| Lumbar Range of Motion, coronal (°) | The angular range of the lumbar spine in the coronal plane (roll) | **×** |  | **×** |
| Lumbar Range of Motion, sagittal (°) | The angular range of the lumbar spine in the sagittal plane (pitch) | **×** |  | **×** |
| Lumbar Range of Motion, transverse (°) | The angular range of the lumbar spine in the transverse plane (yaw) | **×** |  | **×** |
| Harmonic Ratio,  vertical (a.u.) | Harmonic ratio of pelvis linear acceleration in vertical direction, computed as described in methods | **×** |  |  |
| Harmonic Ratio,  antero-posterior (a.u.) | Harmonic ratio of pelvis linear acceleration in antero-posterior direction, computed as described in methods | **×** |  |  |
| Harmonic Ratio,  medio-lateral (a.u.) | Harmonic ratio of pelvis linear acceleration in medio-lateral direction, computed as described in methods | **×** |  |  |
| SPcmp (a.u.) | Spatial variability composite measure, computed from stride length CV and lateral step deviation as describe in methods section and elsewhere (Ilg et al., 2020) | | | |

Definitions of *Mobility Lab* gait measures from https://www.apdm.com/wp-content/uploads/2015/05/02-Mobility-Lab-Whitepaper.pdf

**Supplementary table 2: Individual participant characteristics**

**SPG7 patients**

| **N** | **Sex** | **Age** | **Disease Duration (y)** | **SPRS^mobility^** | **SPRS** | **SARA^PG^** | **SARA** | **FARS-ADL** | **No. strides (LBW)** | **No. strides (SFW)** |
| --- | --- | --- | --- | --- | --- | --- | --- | --- | --- | --- |
| 1 | m | 41-50 | 23 | 9 | 16 | 5 | 12 | 13 | 34 | 341 |
| 2 | f | 51-60 | 35 | 6 | 8 | 4 | 5,5 | 8 | 31 | 449 |
| 3 | m | 41-50 | 18 | 8 | 12 | 4 | 7,5 | 4 | 26 | 331 |
| 4 | f | 61-70 | 11 | 9 | 16 | 4 | 8,5 | 16 | 31 | 291 |
| 5 | m | 61-70 | 16 | 10 | 13 | 2 | 7,5 | 11 | 33 | - |
| 6 | f | 41-50 | 8 | 14 | 23 | 6 | 11,5 | 13,5 | - | 165 |
| 7 | m | 51-60 | 6 | 7 | 15 | 5 | 10,5 | 4 | 25 | - |
| 8 | f | 51-60 | 10 | 10 | 15 | 3 | 7,5 | 9 | 36 | 147 |
| 9 | m | 41-50 | 15 | 8 | 12 | 2 | 10 | 9 | 30 | 179 |
| 10 | m | 51-60 | 18 | 8 | 8 | 7 | 15,5 | 9 | 32 | 298 |
| 11 | m | 41-50 | 2 | 5 | 7 | 2 | 4,5 | 3 | 25 | 287 |
| 12 | f | 61-70 | 11 | 10 | 14 | 4 | 6 | 11 | 35 | 117 |
| 13 | f | 71-80 | 57 | 9 | 15 | 5 | 8,5 | 12 | 27 | 302 |
| 14 | m | 41-50 | 11 | 7 | 11 | 4 | 12 | 7 | 30 | 222 |
| 15 | f | 51-60 | 15 | 12 | 19 | 5 | 12,5 | 13 | 43 | 117 |
| 16 | m | 51-60 | 7 | 10 | 16 | 3 | 7 | 5 | 30 | 210 |
| 17 | m | 51-60 | 15 | 9 | 14 | 3 | 11,5 | 8 | 28 | 202 |
| 18 | m | 41-50 | 11 | 5 | 11 | 3 | 6 | 5 | 22 | 210 |
| 19 | m | 41-50 | 6 | 10 | 17 | 5 | 8 | 7,5 | 28 | 224 |
| 20 | m | 11-20 | 8 | 5 | 6 | 2 | 3,5 | 3 | 27 | 218 |
| 21 | f | 41-50 | 7 | 5 | 7 | 1 | 7,5 | 2 | - | 145 |
| 22 | f | 51-60 | 19 | 8 | 15 | 4 | 7 | 8,5 | 27 | 258 |
| 23 | f | 51-60 | 20 | 14 | 23 | 6 | 9,5 | 10 | 26 | - |
| 24 | m | 51-60 | 13 | 12 | 17 | 6 | 13,5 | 10 | 23 | 109 |
| 25 | m | 41-50 | 16 | 9 | 12 | 4 | 13,5 | 11 | 19 | 55 |
| 26 | m | 61-70 | 26 | 11 | 16 | 6 | 13,5 | 9,5 | 33 | 214 |
| 27 | m | 41-50 | 8 | 13 | 19 | 7 | 13,5 | 15 | 25 | 159 |
| 28 | m | 51-60 | 14 | 6 | 9 | 3 | 8,5 | 3 | 27 | 422 |
| 29 | m | 31-40 | 38 | 8 | 12 | 5 | 13 | 12,5 | 28 | 124 |
| 30 | m | 21-30 | 10 | 6 | 11 | 3 | 9 | 7,5 | 20 | 349 |
| 31 | m | 51-60 | 16 | 8 | 15 | 4 | 6,5 | 6 | 32 | 209 |
| 32 | m | 51-60 | 12 | 11 | 18 | 6 | 9 | 10 | 37 | - |
| 33 | m | 41-50 | 21 | 6 | 8 | 2 | 3,5 | 3 | 27 | 262 |
| 34 | m | 41-50 | 23 | 4 | 6 | 4 | 12 | 10 | - | 367 |
| 35 | m | 31-40 | 22 | 6 | 8 | 4 | 9 | 10,5 | 27 | 367 |
| 36 | m | 41-50 | 23 | 9 | 17 | 5 | 13 | 16 | 33 | 266 |
| 37 | m | 51-60 | 4 | 6 | 10 | 4 | 7 | 12 | 17 | 280 |
| 38 | m | 41-50 | 22 | 15 | 23 | 7 | 18 | 16 | 22 | 57 |
| 39 | m | 41-50 | 15 | 6 | 9 | 1 | 3,5 | 7 | 34 | 252 |
| 40 | m | 31-40 | 10 | 6 | 9 | 2 | 6,5 | 4 | 25 | 255 |
| 41 | f | 51-60 | 17 | 12 | 26 | 5 | 8 | 19 | 31 | 215 |
| 42 | m | 31-40 | 25 | 12 | 12 | 7 | 14,5 | 9 | 23 | 269 |
| 43 | f | 51-60 | 20 | 10 | 18 | 6 | 13 | 13,5 | 36 | - |
| 44 | m | 41-50 | 19 | 6 | 9 | 1 | 6 | 8 | 27 | - |
| 45 | f | 41-50 | 42 | 13 | 19 | 5 | 16 | 17 | 27 | 140 |
| 46 | f | 41-50 | 30 | 8 | 13 | 3 | 8,5 | 5,5 | 33 | - |
| 47 | m | 41-50 | 15 | 9 | 17 | 4 | 9 | 9 | 33 | 180 |
| 48 | m | 61-70 | 39 | 13 | 19 | 5 | 13,5 | 16 | 32 | - |
| 49 | f | 51-60 | 8 | 11 | 24 | 5 | 9 | 7 | 27 | 406 |
| 50 | f | 51-60 | 12 | 13 | 22 | 7 | 8 | 10 | 39 | 180 |
| 51 | m | 51-60 | 10 | 11 | 17 | 5 | 11 | 11 | 28 | - |
| 52 | m | 51-60 | 27 | 3 | 3 | 1 | 7 | 6 | 24 | 173 |
| 53 | f | 51-60 | 12 | 7 | 16 | 2 | 4,5 | 10 | 25 | 372 |
| 54 | f | 61-70 | 8 | 9 | 14 | 4 | 8,5 | 11 | 27 | 316 |
| 55 | m | 41-50 | 9 | 9 | 14 | 3 | 9 | 5 | 29 | 362 |
| 56 | m | 51-60 | 12 | 10 | 15 | 5 | 12 | 8 | 29 | 151 |
| 57 | f | 41-50 | 1 | 7 | 9 | 2 | 3,5 | 2 | 15 | - |
| 58 | m | 61-70 | 5 | 5 | 10 | 2 | 8,5 | 3,5 | 24 | 474 |
| 59 | m | 51-60 | 21 | 9 | 16 | 4 | 8,5 | 5 | 33 | 212 |
| 60 | m | 21-30 | 7 | 7 | 11 | 3 | 10,5 | 9,5 | 33 | 113 |
| 61 | m | 31-40 | 11 | 8 | 13 | 4 | 11 | 9 | 21 | 332 |
| 62 | f | 51-60 | 23 | 14 | 28 | 5 | 11 | 15,5 | 30 | - |
| 63 | m | 61-70 | 3 | 4 | 8 | 3 | 5 | 3,5 | 34 | 111 |
| 64 | m | 51-60 | 10 | 6 | 11 | 2 | 6,5 | 8 | 38 | 256 |
| 65 | f | 61-70 | 7 | 12 | 15 | 5 | 6,5 | 12,5 | 32 | 370 |
| 66 | m | 51-60 | 19 | 7 | 16 | 4 | 9,5 | 6,5 | 26 | 411 |
| 67 | f | 51-60 | 21 | 13 | 22 | 6 | 17 | 15,5 | 40 | - |
| 68 | m | 41-50 | 11 | 6 | 7 | 1 | 9,5 | 4 | - | 160 |
| 69 | m | 41-50 | 16 | 9 | 13 | 5 | 11,5 | 11 | 24 | 182 |

**Healthy controls**

| **N** | **Sex** | **Age** | **SPRS^mobility^** | **SPRS** | **SARA^PG^** | **SARA** | **FARS-ADL** | **No. strides (LBW)** | **No. strides (SFW)** |
| --- | --- | --- | --- | --- | --- | --- | --- | --- | --- |
| 1 | m | 61-70 | 0 | 0 | 0 | 0,5 | 0 | 23 | 407 |
| 2 | m | 21-30 | 2 | 2 | 0 | 0 | 0 | 27 | - |
| 3 | m | 21-30 | 2 | 2 | 0 | 0 | 0 | 28 | - |
| 4 | m | 41-50 | 2 | 2 | 0 | 0 | 0 | 23 | 145 |
| 5 | m | 41-50 | 0 | 0 | 0 | 1 | 1 | 35 | 171 |
| 6 | f | 41-50 | 4 | 4 | 0 | 0 | 0 | 27 | 112 |
| 7 | m | 41-50 | 1 | 1 | 0 | 0,5 | 0 | 25 | 261 |
| 8 | m | 31-40 | 2 | 2 | 0 | 0 | 0 | 27 | 135 |
| 9 | f | 51-60 | 0 | 0 | 0 | 0 | 0 | 37 | - |
| 10 | m | 31-40 | 0 | 0 | 0 | 0,5 | 0 | 23 | 251 |
| 11 | m | 31-40 | 0 | 0 | 0 | 0,5 | 0 | 20 | 210 |
| 12 | m | 21-30 | 0 | 0 | 0 | 0 | 0 | 31 | 279 |
| 13 | f | 41-50 | 0 | 0 | 0 | 1 | 0 | 22 | 219 |
| 14 | f | 71-80 | 2 | 2 | 1 | 4 | 2,5 | 33 | 184 |
| 15 | m | 71-80 | 1 | 1 | 1 | 1 | 0 | 22 | - |
| 16 | f | 61-70 |  |  | 0 | 0 | 0 | 30 | 308 |
| 17 | f | 31-40 | 0 | 0 | 0 | 0 | 0 | 31 | 383 |
| 18 | f | 31-40 | 0 | 0 | 0 | 0 | 0 | 26 | 234 |
| 19 | m | 61-70 | 0 | 0 | 0 | 0,5 | 0 | 29 | 251 |
| 20 | m | 51-60 | 0 | 0 | 0 | 0,5 | 0 | 29 | - |
| 21 | m | 61-70 | 0 | 0 | 0 | 1 | 0 | 20 | 295 |
| 22 | f | 51-60 | 0 | 0 | 0 | 0,5 | 0 | 26 | - |
| 23 | f | 41-50 | 0 | 1 | 0 | 0 | 1 | 28 | 345 |
| 24 | m | 31-40 | 2 | 2 | 0 | 0 | 0 | 26 | - |
| 25 | f | 61-70 | 0 | 0 | 0 | 0 | 0 | 30 | 157 |
| 26 | f | 51-60 | 1 | 1 | 0 | 1 | 0 | 35 | 358 |
| 27 | f | 51-60 | 0 | 0 | 0 | 0 | 0 | 36 | 378 |
| 28 | m | 51-60 | 0 | 2 | 0 | 1 | 0 | - | 124 |
| 29 | m | 71-80 | 0 | 0 | 0 | 3,5 | 0 | 29 | 51 |
| 30 | m | 51-60 | 0 | 1 | 0 | 0 | 0,5 | 29 | - |
| 31 | f | 41-50 | 0 | 0 | 0 | 0 | 0 | 21 | - |
| 32 | m | 61-70 | 0 | 0 | 0 | 1 | 0 | 30 | - |
| 33 | f | 31-40 | 2 | 2 | 0 | 0 | 0 | 29 | - |
| 34 | f | 31-40 | 0 | 0 | 0 | 0 | 0 | 24 | 346 |
| 35 | m | 41-50 | 2 | 2 | 0 | 0 | 0 | 26 | - |
| 36 | f | 61-70 | 0 | 0 | 0 | 0,5 | 0 | 30 | 229 |
| 37 | f | 51-60 | 1 | 1 | 0 | 0 | 0 | 32 | - |
| 38 | f | 31-40 | 0 | 0 | 0 |  | 0 | 26 | 394 |
| 39 | m | 61-70 | 0 | 0 | 0 | 0 | 0 | 22 | 484 |
| 40 | f | 51-60 | 0 | 0 | 0 | 0,5 | 0 | 29 | 385 |
| 41 | f | 51-60 | 0 | 0 | 0 | 0,5 | 0 | 28 | 179 |
| 42 | m | 21-30 | 0 | 0 | 0 | 0 | 0 | 23 | 311 |
| 43 | f | 61-70 | 0 | 0 | 0 | 0 | 0 | 33 | 350 |
| 44 | f | 51-60 | 0 | 0 | 0 | 0 | 0 | 27 | 262 |
| 45 | f | 61-70 | 0 | 0 | 0 | 0,5 | 0 | 26 | 262 |
| 46 | f | 41-50 | 0 | 0 | 0 | 0,5 | 0 | 26 | 228 |
| 47 | f | 41-50 | 2 | 2 | 1 | 1 | 0 | 25 | 352 |
| 48 | m | 41-50 | 0 | 0 | 0 | 0,5 | 0 | 28 | 325 |
| 49 | m | 31-40 | 2 | 2 | 0 | 0 | 0 | 22 | - |
| 50 | f | 41-50 | 1 | 5 | 0 | 0 | 1 | 25 | 324 |
| 51 | f | 21-30 | 0 | 0 | 0 | 0 | 0 | 27 | 327 |

**Supplementary table 3: Discrimination of patients with mild disease severity (SPRS^mobility^ ≤ 9) from HC (lab-based walking)**

|  | **SPG7 (SPRS^mobility^ ≤ 9)** | | **HC** | |  |  |  |
| --- | --- | --- | --- | --- | --- | --- | --- |
| ***N (f/m)*** | 57 (16/41) | | 37 (22/15) | |  |  |  |
| ***Demographic/clinical measures*** | Median | MADN | Median | MADN |  |  |  |
| Age (y) | 48.0 | 10.4 | 47.5 | 17.8 |  |  |  |
| Disease duration (y) | 15.0 | 7.41 |  |  |  |  |  |
| SPRS^mobility^ | 7.00 | 1.48 | 0.00 | 0.00 |  |  |  |
| SPRS | 12.0 | 4.45 | 0.00 | 0.00 |  |  |  |
| SARA^PG^ | 4.00 | 1.48 | 0.00 | 0.00 |  |  |  |
| SARA | 8.50 | 2.97 | 0.00 | 0.00 |  |  |  |
| FARS-ADL | 8.00 | 4.45 | 0.00 | 0.00 |  |  |  |
| ***Gait measures*** | Median | MADN | Median | MADN | Cliff’s δ | p-value |  |
| SPcmp | 0.319 | 0.0905 | 0.158 | 0.0659 | 0.86 | 2.3e-12 | * |
| Swing CV | 0.0196 | 0.0062 | 0.0121 | 0.0034 | 0.80 | 6.3e-11 | * |
| Lateral Step Deviation (%) | 3.18 | 0.846 | 1.94 | 0.515 | 0.78 | 1.5e-10 | * |
| Pitch at Initial Contact (°) | 2.10 | 0.587 | 3.63 | 0.835 | -0.77 | 3.7e-10 | * |
| Harmonic Ratio V | 19.8 | 5.30 | 27.5 | 4.83 | -0.75 | 3.8e-09 | * |
| Harmonic Ratio AP | 2.02 | 0.465 | 3.19 | 0.913 | -0.70 | 9.1e-09 | * |
| Pitch at Toe Off (°) | 32.9 | 5.11 | 37.9 | 4.05 | -0.68 | 2.3e-08 | * |
| Stride Time CV | 1.40 | 0.394 | 0.887 | 0.219 | 0.67 | 3.8e-08 | * |
| Circumduction | 4.64 | 1.64 | 2.79 | 0.948 | 0.65 | 1.3e-07 | * |
| Double Support MADN | 1.79 | 0.473 | 2.47 | 0.638 | -0.62 | 4.2e-07 | * |
| Stride Length (m) | 0.0248 | 0.0081 | 0.0169 | 0.0067 | 0.61 | 6.1e-07 | * |
| Gait Speed (m/s) | 1.13 | 0.127 | 1.33 | 0.185 | -0.59 | 1.3e-06 | * |
| Harmonic Ratio ML | 1.23 | 0.547 | 0.791 | 0.295 | 0.56 | 4.8e-06 | * |
| Pitch at Toe Off MADN | 1.00 | 0.182 | 1.24 | 0.203 | -0.55 | 6.7e-06 | * |
| Stride Length CV | 24.6 | 4.54 | 19.5 | 2.72 | 0.54 | 9.7e-06 | * |
| Double Support (%) | 37.7 | 2.36 | 40.3 | 1.38 | -0.53 | 1.3e-05 | * |
| Swing (%) | 10.9 | 3.99 | 15.1 | 3.82 | -0.50 | 3.8e-05 | * |
| Pitch at Initial Contact MADN | 0.0361 | 0.0166 | 0.0255 | 0.0074 | 0.50 | 5.2e-05 | * |
| LRoM transverse (°) | 12.4 | 4.72 | 8.94 | 2.70 | 0.45 | 0.00027 | * |
| Toe Out Angle MADN | 1.46 | 0.525 | 1.05 | 0.363 | 0.41 | 0.0012 | * |
| Elevation at Midswing MADN | 1.89 | 0.660 | 1.47 | 0.660 | 0.40 | 0.0012 | * |
| Pitch at Mid Swing (°) | 0.301 | 0.129 | 0.235 | 0.0666 | 0.39 | 0.0015 | * |
| Stride Time (s) | 1.75 | 0.950 | 1.18 | 0.480 | 0.38 | 0.0018 |  |
| LRoM sagittal (°) | 6.18 | 2.39 | 4.78 | 1.42 | 0.32 | 0.01 |  |
| LRoM sagittal CV | 0.180 | 0.0553 | 0.138 | 0.0423 | 0.32 | 0.01 |  |
| Elevation at Midswing (cm) | 1.13 | 0.0973 | 1.07 | 0.0620 | 0.26 | 0.031 |  |
| Toe Out Angle (°) | 9.02 | 4.91 | 8.02 | 5.53 | 0.13 | 0.29 |  |
| LRoM coronal CV | 7.33 | 2.28 | 7.69 | 2.18 | -0.07 | 0.55 |  |
| LRoM coronal (°) | 0.191 | 0.0681 | 0.184 | 0.0711 | 0.06 | 0.64 |  |
| LRoM transverse CV | 0.0800 | 0.0251 | 0.0896 | 0.0401 | -0.02 | 0.9 |  |

**Supplementary table 4: Correlation of gait measures with exploratory clinical measures (all patients, lab-based walking)**

|  | **SPRS** | | **SARA** | | **FARS-ADL** | |
| --- | --- | --- | --- | --- | --- | --- |
| SPcmp | **0.61** | **[0.44, 0.73] *** | **0.46** | **[0.24, 0.64] *** | **0.39** | **[0.18, 0.56] *** |
| Stride Time CV | **0.56** | **[0.35, 0.71] *** | **0.53** | **[0.33, 0.69] *** | **0.38** | **[0.15, 0.56] *** |
| Swing CV | **0.50** | **[0.28, 0.66] *** | **0.51** | **[0.26, 0.68] *** | **0.44** | **[0.22, 0.61] *** |
| Stride Length CV | **0.55** | **[0.36, 0.70] *** | **0.57** | **[0.37, 0.71] *** | **0.45** | **[0.23, 0.62] *** |
| Stride Length (m) | **-0.52** | **[-0.67, -0.34] *** | **-0.30** | **[-0.52, -0.05] *** | **-0.38** | **[-0.56, -0.14] *** |
| Gait Speed (m/s) | **-0.50** | **[-0.66, -0.28] *** | **-0.30** | **[-0.50, -0.04] *** | **-0.41** | **[-0.59, -0.18] *** |
| Pitch at Toe Off (°) | **-0.54** | **[-0.69, -0.37] *** | **-0.41** | **[-0.61, -0.14] *** | **-0.35** | **[-0.54, -0.12] *** |
| Lateral Step Deviation (%) | **0.41** | **[0.15, 0.61] *** | **0.36** | **[0.10, 0.56] *** | 0.21 | [-0.05, 0.44] |
| Double Support (%) | **0.51** | **[0.31, 0.66] *** | 0.17 | [-0.08, 0.39] | **0.35** | **[0.12, 0.52] *** |
| Swing (%) | **-0.50** | **[-0.68, -0.32] *** | -0.17 | [-0.40, 0.08] | **-0.34** | **[-0.51, -0.14] *** |
| Double Support MADN | **0.36** | **[0.13, 0.56] *** | **0.44** | **[0.23, 0.60] *** | **0.34** | **[0.13, 0.52] *** |
| Pitch at Initial Contact (°) | **-0.35** | **[-0.57, -0.08] *** | -0.19 | [-0.42, 0.07] | -0.14 | [-0.42, 0.14] |
| Pitch at Toe Off MADN | 0.21 | [-0.04, 0.45] | **0.48** | **[0.27, 0.64] *** | **0.26** | **[0.04, 0.47] *** |
| Pitch at Initial Contact MADN | **0.28** | **[0.04, 0.51] *** | **0.37** | **[0.13, 0.58] *** | 0.19 | [-0.06, 0.38] |
| Harmonic Ratio AP | -0.17 | [-0.39, 0.07] | -0.16 | [-0.39, 0.08] | -0.17 | [-0.40, 0.10] |
| Circumduction | **0.41** | **[0.19, 0.58] *** | 0.23 | [-0.02, 0.45] | 0.09 | [-0.14, 0.31] |
| Harmonic Ratio V | -0.20 | [-0.41, 0.04] | **-0.29** | **[-0.48, -0.04] *** | -0.06 | [-0.30, 0.18] |
| Harmonic Ratio ML | -0.06 | [-0.28, 0.17] | -0.16 | [-0.39, 0.07] | -0.15 | [-0.37, 0.11] |

Spearman’s ρ [95% CI]. *: p<0.05

**Supplementary table 5: Correlation of gait measures with exploratory clinical measures (mild patient cohort, lab-based walking)**

|  | **SPRS** | | **SARA** | | **FARS-ADL** | |
| --- | --- | --- | --- | --- | --- | --- |
| Swing CV | 0.22 | [-0.11, 0.50] | **0.33** | **[0.00, 0.62] *** | **0.36** | **[0.03, 0.61] *** |
| SPcmp | **0.36** | **[0.08, 0.58] *** | 0.26 | [-0.06, 0.53] | 0.29 | [-0.03, 0.54] |
| Pitch at Toe Off MADN | 0.19 | [-0.15, 0.49] | **0.51** | **[0.26, 0.72] *** | **0.32** | **[0.02, 0.58] *** |
| Stride Time CV | 0.24 | [-0.12, 0.52] | **0.37** | **[0.09, 0.58] *** | 0.10 | [-0.21, 0.42] |
| Double Support MADN | 0.20 | [-0.09, 0.51] | 0.28 | [-0.03, 0.54] | 0.23 | [-0.09, 0.50] |
| Pitch at Toe Off (°) | **-0.32** | **[-0.55, -0.04] *** | -0.29 | [-0.56, 0.03] | -0.07 | [-0.38, 0.25] |
| Stride Length (m) | -0.11 | [-0.39, 0.19] | -0.15 | [-0.43, 0.17] | -0.03 | [-0.37, 0.29] |
| Gait Speed (m/s) | -0.12 | [-0.41, 0.21] | -0.06 | [-0.35, 0.23] | -0.05 | [-0.37, 0.25] |
| Swing (%) | -0.28 | [-0.52, 0.01] | -0.02 | [-0.35, 0.31] | -0.04 | [-0.32, 0.27] |
| Double Support (%) | 0.26 | [-0.02, 0.51] | -0.00 | [-0.32, 0.34] | 0.01 | [-0.28, 0.28] |
| Lateral Step Deviation (%) | 0.15 | [-0.22, 0.43] | 0.02 | [-0.34, 0.34] | -0.06 | [-0.40, 0.27] |
| Pitch at Mid Swing (°) | -0.26 | [-0.51, 0.05] | -0.06 | [-0.36, 0.25] | -0.01 | [-0.33, 0.32] |
| Harmonic Ratio ML | 0.03 | [-0.27, 0.31] | -0.08 | [-0.37, 0.17] | 0.04 | [-0.27, 0.35] |
| Pitch at Initial Contact (°) | -0.04 | [-0.34, 0.28] | -0.01 | [-0.31, 0.32] | -0.03 | [-0.36, 0.34] |
| Circumduction | 0.23 | [-0.08, 0.50] | 0.06 | [-0.29, 0.40] | -0.11 | [-0.38, 0.22] |
| Harmonic Ratio V | 0.05 | [-0.22, 0.34] | -0.12 | [-0.40, 0.18] | 0.15 | [-0.13, 0.44] |
| Harmonic Ratio AP | 0.10 | [-0.26, 0.39] | -0.04 | [-0.35, 0.28] | -0.01 | [-0.32, 0.29] |

Spearman’s ρ [95% CI]. *: p < 0.05

**Supplementary table 6: Correlations between lab-based walking (LBW) and supervised free walking (SFW) conditions**

| **Gait measure** | **Spearman’s ρ [95% CI]** | |
| --- | --- | --- |
| Pitch at Toe Off (°) | 0.93 | [0.90, 0.96] |
| Pitch at Initial Contact (°) | 0.93 | [0.87, 0.97] |
| Pitch at Mid Swing (°) | 0.90 | [0.81, 0.95] |
| Stride Length (m) | 0.90 | [0.80, 0.95] |
| LRoM transverse (°) | 0.87 | [0.77, 0.93] |
| Toe Out Angle (°) | 0.86 | [0.71, 0.94] |
| LRoM coronal (°) | 0.85 | [0.73, 0.93] |
| Circumduction | 0.85 | [0.65, 0.94] |
| Swing (%) | 0.85 | [0.70, 0.92] |
| Double Support (%) | 0.84 | [0.68, 0.92] |
| Harmonic Ratio AP | 0.82 | [0.67, 0.91] |
| LRoM sagittal (°) | 0.81 | [0.66, 0.90] |
| Harmonic Ratio V | 0.80 | [0.61, 0.91] |
| Gait Speed (m/s) | 0.80 | [0.65, 0.89] |
| Elevation at Midswing (cm) | 0.79 | [0.63, 0.89] |
| Stride Time (s) | 0.75 | [0.49, 0.88] |
| Harmonic Ratio ML | 0.74 | [0.56, 0.84] |
| LRoM coronal CV | 0.68 | [0.53, 0.79] |
| Swing CV | 0.65 | [0.40, 0.81] |
| LRoM transverse CV | 0.62 | [0.37, 0.78] |
| Toe Out Angle MADN | 0.56 | [0.30, 0.75] |
| Elevation at Midswing MADN | 0.56 | [0.33, 0.72] |
| Stride Time CV | 0.55 | [0.29, 0.73] |
| Double Support MADN | 0.53 | [0.28, 0.72] |
| Lateral Step Deviation (%) | 0.53 | [0.27, 0.72] |
| Stride Length CV | 0.53 | [0.27, 0.71] |
| LRoM sagittal CV | 0.51 | [0.27, 0.68] |
| SPcmp | 0.47 | [0.20, 0.68] |
| Pitch at Toe Off MADN | 0.43 | [0.16, 0.64] |
| Pitch at Initial Contact MADN | 0.38 | [0.08, 0.62] |

**Supplementary table 7: Correlation of gait measures with exploratory clinical measures (all patients, supervised free walking)**

|  | **SPRS^mobility^** | | **SARA^PG^** | | **SPRS** | | **SARA** | | **FARS-ADL** | |
| --- | --- | --- | --- | --- | --- | --- | --- | --- | --- | --- |
| Gait Speed (m/s) | **-0.59** | **[-0.76, -0.35] **** | -0.32 | [-0.55, -0.03] * | **-0.45** | **[-0.66, -0.18] **** | **-0.26** | **[-0.51, 0.04] *** | **-0.39** | **[-0.61, -0.10] *** |
| Stride Time CV | **0.57** | **[0.29, 0.73] **** | 0.38 | [0.10, 0.60] * | **0.46** | **[0.20, 0.65] **** | **0.35** | **[0.05, 0.60] *** | **0.27** | **[0.00, 0.50] *** |
| Stride Length (m) | **-0.55** | **[-0.71, -0.31] **** | -0.28 | [-0.52, 0.02] * | **-0.42** | **[-0.62, -0.15] **** | -0.19 | [-0.44, 0.11] | **-0.27** | **[-0.51, 0.01] *** |
| Stride Length CV | **0.54** | **[0.26, 0.71] **** | **0.47** | **[0.17, 0.66] **** | **0.39** | **[0.09, 0.62] *** | **0.48** | **[0.21, 0.69] **** | 0.25 | [-0.04, 0.49] |
| Double Support MADN | **0.52** | **[0.28, 0.71] **** | **0.46** | **[0.15, 0.65] **** | **0.37** | **[0.12, 0.59] *** | **0.53** | **[0.29, 0.71] **** | **0.32** | **[0.08, 0.53] *** |
| Pitch at Initial Contact (°) | **-0.51** | **[-0.69, -0.28] **** | -0.36 | [-0.58, -0.07] * | **-0.44** | **[-0.63, -0.19] **** | -0.25 | [-0.48, 0.02] | -0.17 | [-0.42, 0.11] |
| Swing CV | **0.51** | **[0.26, 0.69] **** | 0.40 | [0.12, 0.61] * | **0.35** | **[0.09, 0.58] *** | **0.44** | **[0.15, 0.66] **** | **0.28** | **[0.04, 0.50] *** |
| Pitch at Toe Off (°) | **-0.46** | **[-0.65, -0.22] **** | -0.27 | [-0.54, 0.05] * | **-0.43** | **[-0.63, -0.21] **** | -0.22 | [-0.51, 0.07] | -0.20 | [-0.44, 0.07] |
| Pitch at Toe Off MADN | **0.45** | **[0.21, 0.65] **** | **0.46** | **[0.20, 0.66] **** | 0.24 | [-0.05, 0.48] | **0.50** | **[0.24, 0.70] **** | **0.29** | **[0.00, 0.52] *** |
| SPcmp | **0.43** | **[0.15, 0.63] **** | 0.34 | [0.03, 0.58] * | **0.28** | **[0.00, 0.52] *** | **0.27** | **[-0.01, 0.51] *** | 0.24 | [-0.05, 0.50] |
| Lateral Step Deviation (%) | 0.37 | [0.09, 0.59] * | 0.30 | [0.00, 0.53] * | 0.23 | [-0.06, 0.48] | 0.19 | [-0.09, 0.44] | 0.23 | [-0.05, 0.47] |
| Circumduction | 0.30 | [0.06, 0.51] * | 0.05 | [-0.24, 0.31] | 0.32 | [0.08, 0.53] * | 0.12 | [-0.17, 0.37] | -0.01 | [-0.27, 0.25] |
| Double Support (%) | 0.28 | [-0.02, 0.51] * | 0.03 | [-0.27, 0.32] | **0.31** | **[0.03, 0.55] *** | -0.06 | [-0.34, 0.25] | 0.15 | [-0.11, 0.38] |
| Harmonic Ratio AP | -0.28 | [-0.49, -0.03] * | -0.36 | [-0.56, -0.11] * | -0.13 | [-0.38, 0.11] | **-0.26** | **[-0.50, -0.01] *** | -0.15 | [-0.40, 0.11] |
| Swing (%) | -0.28 | [-0.52, 0.01] * | -0.02 | [-0.31, 0.29] | **-0.31** | **[-0.55, -0.04] *** | 0.07 | [-0.23, 0.33] | -0.15 | [-0.39, 0.11] |
| Harmonic Ratio V | -0.15 | [-0.39, 0.11] | -0.11 | [-0.39, 0.16] | -0.03 | [-0.26, 0.20] | -0.23 | [-0.49, 0.05] | -0.02 | [-0.25, 0.25] |
| Harmonic Ratio ML | -0.08 | [-0.34, 0.19] | -0.12 | [-0.37, 0.12] | 0.10 | [-0.16, 0.35] | -0.13 | [-0.39, 0.14] | -0.10 | [-0.34, 0.18] |

Spearman’s ρ [95% CI]. **: p < 0.05 / (2 x 17); *: p < 0.05
