## supplementary_figures for "Towards patient-relevant, trial-ready digital motor outcomes for SPG7: a cross-sectional prospective multi-center study (PROSPAX)"

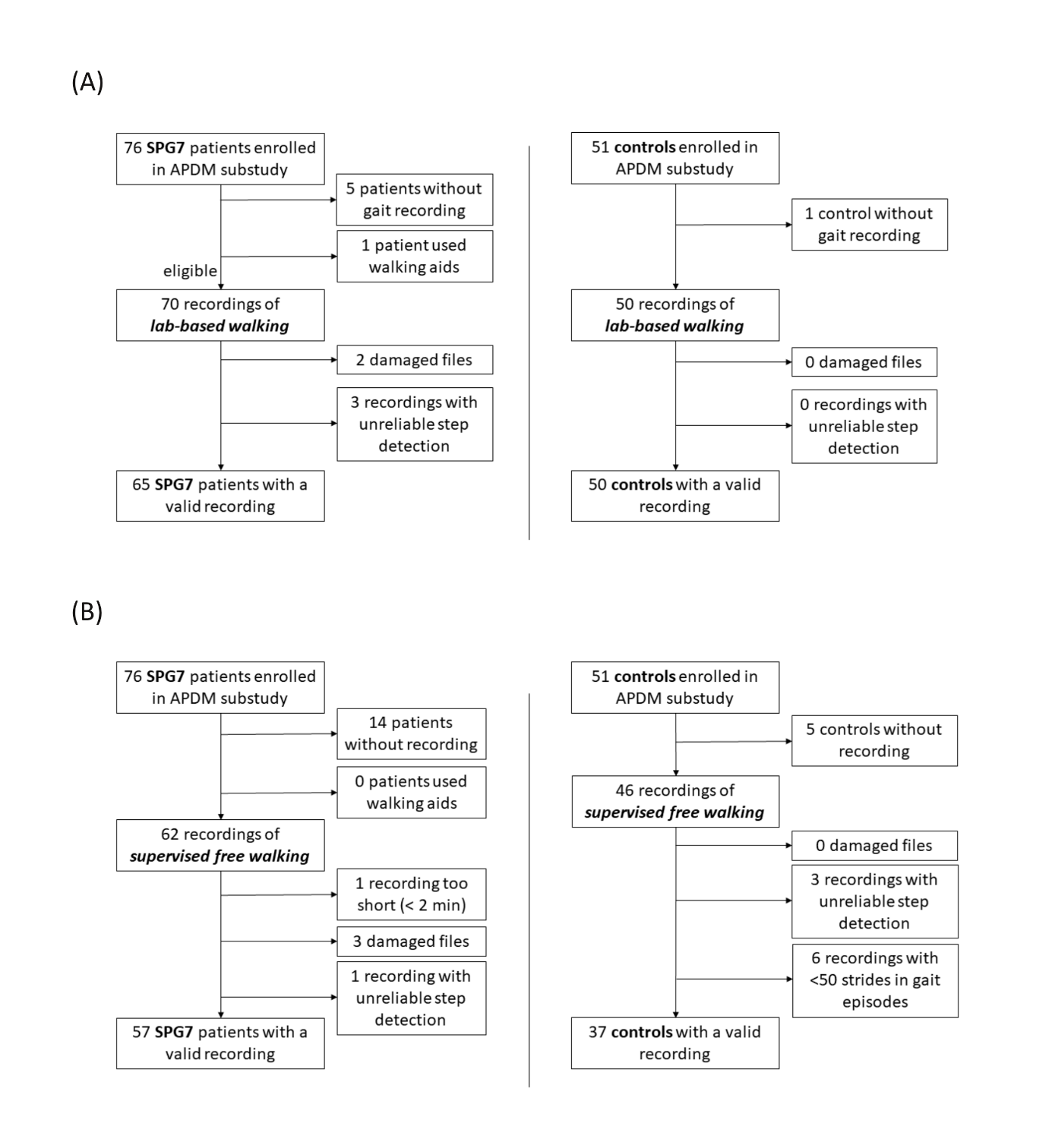
**Supplementary figure 1**: Inclusion and exclusion of gait recordings from SPG7 patients (left) and healthy controls (right) for the lab-based walking (A) and supervised free walking (B) conditions.

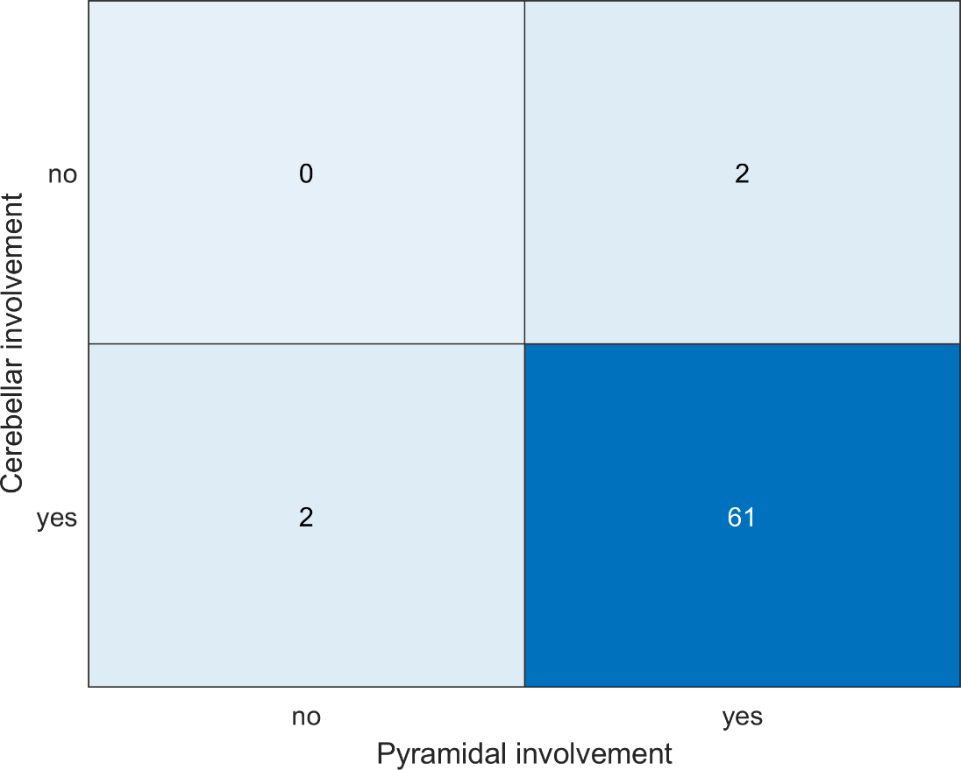

**Supplementary figure 2:** To investigate, whether SPG7 patients with purely spastic or purely ataxic phenotype were included in the study, cerebellar and pyramidal systems involvement was determined by adding relevant clinical items as follows. Scores of ≥ 1 were rated as presence of cerebellar or pyramidal involvement, respectively:

- Cerebellar involvement = SARA items 4-7: speech, finger-nose, finger-chase, alternating hand movements + INAS items: hypometric / hypermetric saccades + horizontal / vertical GEN
- Pyramidal involvement = SPRS items 7-8: spasticity of hip adductors and upon knee flexion + INAS: spasticity of gait, upper limbs, lower limbs + INAS: hyperreflexia of biceps, patellar and achilles reflexes + INAS: presence of extensor plantar reflex

GEN: gaze-evoked nystagmus. INAS: Inventory of Non-Ataxia Signs. SARA: Scale for the Assessment and Rating of Ataxia. SPRS: Spastic Paraplegia Rating Scale.

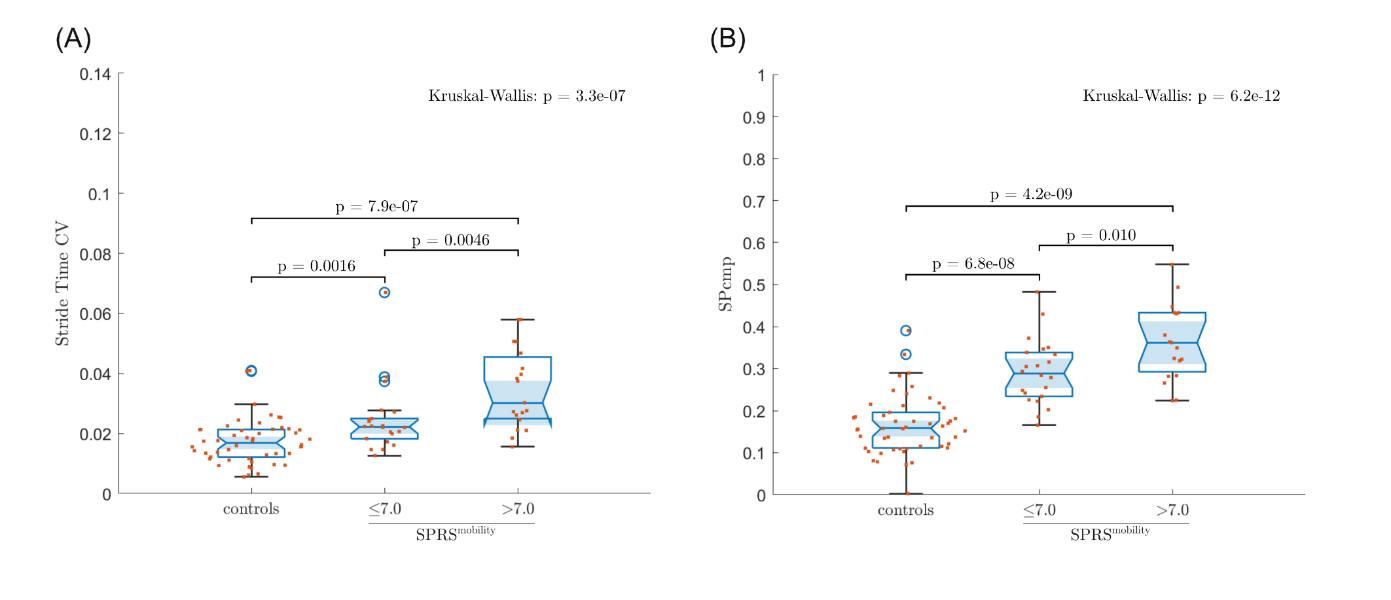

**Supplementary figure 3:** As an alternative illustration of the associations between gait measures and the SPRS^mobility^ within the mild patient cohort, the mild patient subgroup was further subdivided by again performing a median split with respect to SPRS^mobility^. Between-group differences were analysed using the Kruskal-Wallis test. When the Kruskal-Wallis test yielded a significant effect, post hoc analysis was performed using a Wilcoxon ranksum test. To correct for number of measures (=30) analysed, Kruskal-Wallis test and Wilcoxon ranksum test were considered significant when p < 0.05/30.

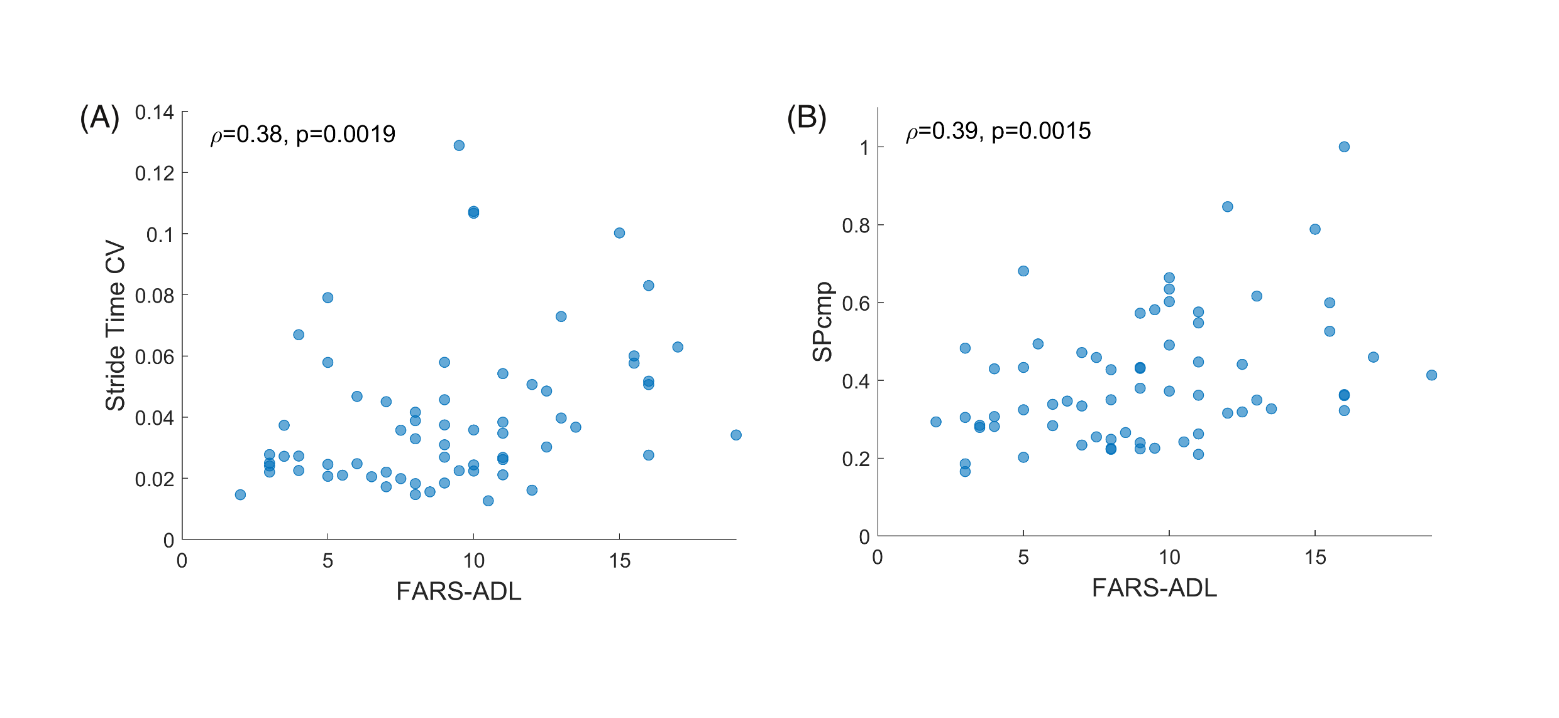

**Supplementary figure 4:** Gait measures Stride Time CV (A) and SPcmp (B) plotted against the activities of daily living subscore of the Friedreich Ataxia Rating Scale (FARS-ADL) for all SPG7 patients in the lab-based walking condition. Effect size of correlation Spearman’s ρ and associated p value given in upper left corners.
